# The Impact of Evolving Endometriosis Guidelines on Diagnosis and Observational Health Studies

**DOI:** 10.1101/2024.12.13.24319010

**Authors:** Harry Reyes Nieva, Aparajita Kashyap, Erica A. Voss, Anna Ostropolets, Adit Anand, Mert Ketenci, Frank J. Defalco, Young Sang Choi, Yanwei Li, Monica N. Allen, Stephanie A. Guang, Karthik Natarajan, Patrick Ryan, Noémie Elhadad

## Abstract

**STUDY QUESTION:** Do recent changes in European Society of Human Reproduction and Embryology (ESHRE) clinical guidelines result in more comprehensive diagnosis of women with endometriosis?

**SUMMARY ANSWER:** The latest shift in clinical guidelines results in diagnosis of more women with endometriosis but current ESHRE diagnostic criteria do not capture a sizable percentage of women with the disease.

**WHAT IS KNOWN ALREADY:** Historically, laparoscopy was the gold standard for diagnosing endometriosis, a complex gynecological condition marked by a heterogeneous set of symptoms that vary widely among women. More recently, changes in clinical guidelines have shifted to incorporate imaging-based approaches such as transvaginal sonography and magnetic resonance imaging.

**STUDY DESIGN, SIZE, DURATION:** Retrospective, observational cohort study of women aged 15-49 years diagnosed with endometriosis in the United States (US) between January 1, 2013, and December 31, 2023.

**PARTICIPANTS/MATERIALS, SETTING, METHODS:** Data sources include US insurance claims data from the Merative™ MarketScan® Commercial Database (CCAE), Merative™ MarketScan® Multi-State Medicaid Database (MDCD), Optum® de-identified Electronic Health Record dataset (Optum® EHR), and electronic health record (EHR) data from a large academic medical center in New York City (CUIMC EHR). To examine the potential impact of expanding clinical criteria for the diagnosis of endometriosis, we validated and compared five cohort definitions based on different sets of diagnostic guidelines involving combinations of surgical confirmation, diagnostic imaging, guideline-recognized symptoms, and other symptoms commonly reported among women with endometriosis. We performed pairwise comparisons between cohorts and applied Bonferroni corrections to account for multiple comparisons.

**MAIN RESULTS AND THE ROLE OF CHANCE:** We identified 491,048 women with a diagnosis of endometriosis across the CCAE, MDCD, Optum EHR, and CUIMC EHR datasets. Each cohort definition demonstrated strong positive predictive value (0.84-0.96), yet only 15-20% of cases were identified by all 5 cohort definitions. Women diagnosed with endometriosis based on imaging and symptoms were three years younger, on average, than women with a diagnosis based on surgical confirmation (mean age = 35 years [SD = 9] vs 38 years [SD = 8]; p<0.001). Women in cohorts based only on symptoms were two years younger than those based on surgery (36 years [SD = 8] vs 38 years [SD = 8]; p<0.001). More than one-fourth of cases presented with endometriosis-related symptoms but lacked surgical or imaging-related documentation required by ESHRE guideline criteria. Pain was reported among nearly all women with endometriosis. Abdominal pain and pain in the pelvis were the most prevalent (ranging from 69% to 90% of women in each cohort). Among approximately 2-5% of all endometriosis cases (14,795 total), women presented with pelvic and/or abdominal pain but none of the other symptoms noted in clinical guidelines.

**LIMITATIONS, REASONS FOR CAUTION:** Our study has potential biases associated with documentation practices and secondary data use of insurance claims and EHR data. Further, the phenotyping algorithms used rely on clinical codes that do not necessarily capture all ESHRE diagnostic criteria for endometriosis and may not be generalizable to women with atypical presentation of endometriosis.

**WIDER IMPLICATIONS OF THE FINDINGS:** High positive predictive value among all five cohort definitions despite poor overlap among cases identified illustrates both the heterogeneous presentation of the disease and importance of expanding diagnostic criteria. For example, cohorts derived from updated guidelines identified younger patients at time of diagnosis. Women diagnosed based on imaging had higher rates of emergency room visits while patients diagnosed via laparoscopy had a larger number of hospitalizations. The substantial number of cases with pelvic and/or abdominal pain but none of the other symptoms noted in clinical guidelines underscores the continued need for improved access to timely and appropriate care, particularly among those with non-classical symptoms, different care-seeking patterns, or lack of available surgical intervention.

## INTRODUCTION

Endometriosis is a chronic gynecological condition characterized by the growth of endometrial tissue outside of the uterus.^1,2^ Marked by a heterogeneous set of symptoms that vary widely among women, the complex disorder can impact a range of body systems and often involves chronic pain, dysmenorrhea, dyspareunia, dysuria, and fatigue. Half of women with infertility are diagnosed with endometriosis.^1,2^ Given the enigmatic nature of the disease and high likelihood for underdiagnosis, estimates of prevalence can vary widely. Nonetheless, the statistic most often cited is that 10% of women have endometriosis.^3^

Historically, laparoscopy was the gold standard for diagnosing endometriosis.^4^ While effective in confirming the presence of the disease, laparoscopy poses potential risks and is not always preferred by patients. The correlation between the extent of surgical findings and severity of symptoms also remains complex, underscoring the need for a comprehensive diagnostic strategy. Additionally, there is growing concern that emphasis on laparoscopy contributes to delays and underdiagnosis, particularly in cases where surgical intervention is not feasible or not covered by insurance. Notably, the average delay between disease onset and formal diagnosis of endometriosis is considered to be between four and eleven years.^5–7^

In 2022 the European Society of Human Reproduction and Embryology (ESHRE) shifted its clinical guidelines regarding endometriosis toward a more multimodal approach to diagnosis, emphasizing assessment of indicative symptoms and the use of diagnostic imaging such as transvaginal sonography (TVS) and magnetic resonance imaging (MRI) as a complement to laparoscopic confirmation.^8,9^ The shift in clinical guidelines acknowledges high variability in diagnosing endometriosis and aims to enhance accuracy and expedite diagnosis.

Given the potential for reducing delayed and underdiagnosis, the rise of new guidelines also suggests prior characterizations may not accurately reflect the full composition, care patterns, and spectrum of experiences associated with endometriosis, especially prior to receipt of a formal diagnosis. It is not yet clear how the expansion of diagnostic criteria will impact the composition of women diagnosed with the disorder. Moreover, these new guidelines may still not reflect the breadth of symptoms commonly experienced by women with endometriosis. Importantly, it is critical to investigate how such changes and considerations are likely to influence future observational health research given the impact on both clinical practice and documentation of care.

In this study, we sought to examine differences among women diagnosed with endometriosis, including their clinical histories and demographics. Specifically, we compared five cohort definitions based on different sets of diagnostic criteria involving combinations of surgical confirmation, diagnostic imaging, guideline-recognized symptoms, and other symptoms commonly reported among women with endometriosis.

## MATERIALS AND METHODS

### Study design, setting, and population

We performed a retrospective, observational cohort study of women in the United States (US) aged 15-49 years with a diagnosis date of endometriosis between January 1, 2013 and December 31, 2023. This study was approved by the institutional review board of Columbia University Irving Medical Center (Protocol AAAO7805) and reporting follows the Strengthening the Reporting of Observational Studies in Epidemiology (STROBE) guideline.

### Data sources

Our data sources included US insurance claims from the Merative™ MarketScan® Commercial Database (CCAE) and Merative™ MarketScan® Multi-State Medicaid (MDCD) databases as well as administrative and electronic health record (EHR) data from Optum® de-identified EHR (Optum® EHR) and Columbia University Irving Medical Center (CUIMC) databases. To standardize the data, each de-identified dataset was transformed to the Observational Health Data Sciences and Informatics (OHDSI) Observational Medical Outcomes Partnership (OMOP) Common Data Model (CDM).^10,11^

The CCAE dataset includes health insurance claims across the care continuum (i.e., inpatient, outpatient, outpatient pharmacy, and carve-out behavioral healthcare) as well as enrollment data from large employers and health plans across the US who provide private healthcare coverage for more than 170 million employees, their spouses, and dependents. This administrative claims database includes a variety of fee-for-service, preferred provider organizations, and capitated health plans.

The MDCD dataset reflects the healthcare service use of more than 35 million individuals covered by Medicaid programs in numerous geographically dispersed states. The database contains the pooled healthcare experience of Medicaid enrollees, covered under fee-for-service and managed care plans. It includes records of inpatient services, inpatient admissions, outpatient services, and prescription drug claims, as well as information on long-term care.

The Optum® EHR dataset contains clinical, claims, and other administrative data on more than 111 million patients from more than 111,000 sites of care spanning dozens of healthcare provider organizations across the US. Data are obtained from inpatient and ambulatory EHRs, practice management systems, and other internal systems. Data elements include demographics, prescribed and administered medications, immunizations, allergies, laboratory results, vital signs and other observable measurements, clinical and inpatient stay administrative data and coded diagnoses and procedures.

The CUIMC EHR dataset comprises 6.7 million patients with data collection starting in 1985. CUIMC is a quaternary care center in the northeast U.S. with primary care practices in northern Manhattan and surrounding areas. The CUIMC database currently holds information about patient demographics, conditions, symptoms and other observations, laboratory and vital sign measurements, medications, medical devices, and both inpatient and outpatient encounters.

### Cohort definitions

Based on a combination of coded diagnoses, observations, procedures, and measurements, we constructed five different cohorts — (i) women who received a diagnosis of endometriosis based on surgical confirmation (Cohort A); (ii) women who received a diagnosis of endometriosis based on a combination of diagnostic imaging and presentation of guideline-recognized endometriosis symptoms (Cohort B); (iii) women who received a diagnosis of endometriosis and presented with guideline-recognized endometriosis symptoms, regardless of whether diagnostic imaging was performed (Cohort C); (iv) women who received a diagnosis of endometriosis and presented with guideline-recognized endometriosis symptoms and/or pelvic pain (Cohort D); and (v) women who received a diagnosis of endometriosis and presented with guideline-recognized endometriosis symptoms, pelvic pain, and/or abdominal pain (Cohort E). For each cohort, we restricted our analysis to participants with at least one year of continuous observation prior to cohort entry to ensure we capture incident events. Each cohort definition was translated into a phenotype,^12^ a computable algorithm incorporating temporal logic, to query the CCAE, MDCD, Optum® EHR and CUIMC databases.

#### Women with endometriosis diagnosis based on surgery

For Cohort A, women with an endometriosis-related surgical procedure (e.g., laparoscopic surgery), we required an accompanying endometriosis diagnosis within 30 days of the procedure. For this group, aligning with previous guidelines, the date of cohort entry was the date of surgery.

#### Women with endometriosis diagnosis based on imaging and guideline-recognized symptoms

Based on the updated diagnosis guidelines, our definition for Cohort B, women with an endometriosis diagnosis based on imaging required the occurrence of (i) at least one medical imaging code (e.g., transvaginal ultrasound); (ii) at least two endometriosis diagnosis codes on the date of or following imaging; (iii) at least one code for dysmenorrhea, dyspareunia, or dysuria prior to diagnosis; and (iv) at least one other code for an endometriosis-related symptom prior to diagnosis, including additional symptoms such as constipation and cough to account for episodes of dyschezia and cyclical cough, respectively, which are known indications of endometriosis. Due to data limitations, more granular symptom descriptors were not available in the source data. Additionally, cyclical scar swelling and pain was the only symptom from the updated guidelines without an associated code in the source data and thus not included in the phenotype. For this group, the date of cohort entry was the date of diagnosis.

#### Women with endometriosis diagnosis based only on symptoms

For women who received a diagnosis of endometriosis and presented with endometriosis-related symptoms, we created three cohorts by successively expanding the number of potential symptoms included. For Cohort C, we required the occurrence of (i) at least two endometriosis diagnosis codes; (ii) at least one code for dysmenorrhea, dyspareunia, or dysuria prior to diagnosis; and (iii) at least one other code for an endometriosis-related symptom recognized in the clinical guidelines prior to diagnosis. For Cohort D, we required the same criteria as Cohort C but included pelvic pain. Cohort E builds upon Cohort D further by including abdominal pain.

We chose to include Cohorts D and E in our analysis based on patient-reported outcomes compiled via the Phendo mobile app (https://citizenendo.org/phendo/). Phendo is a research-oriented app that has 17,000+ users across the world who submit information about their experiences with endometriosis. Approximately 60% of Phendo users report experiencing chronic pelvic pain and/or abdominal pain.

To validate and compare the relative performance of our five endometriosis cohort definitions, two physicians specializing in gynecology (MNA, SG) independently conducted a detailed chart review of CUIMC EHRs for a sample of patients. Full details regarding phenotype evaluation are provided in Supplementary Materials. In summary, all five cohort definitions achieved high levels of positive predictive value (PPV range: 0.84-0.96) but there was poor concordance among the patients identified in each cohort. Complete agreement among all five cohort definitions was only found among 16% of cases (Fleiss’ kappa = 0.13).

### Outcomes and variables

The primary outcomes of interest were the age at diagnosis, the number and proportion of patients shared among and unique to each cohort, and the prevalence of conditions, medications, and procedures documented prior to receipt of an endometriosis diagnosis. We standardized medications based on their anatomical therapeutic chemical (ATC) level three classification. Secondary outcomes included measures of healthcare utilization including the number of clinical encounters and emergency department visits and/or inpatient hospitalizations occurring before diagnosis. Among participants identified by multiple cohort definitions, we also assessed differences in their date of entry to each cohort.

### Statistical analysis

We calculated descriptive statistics using count, percentage, mean, median, standard deviation, and interquartile range (IQR). We performed pairwise comparisons of proportion between cohorts using Chi-squared tests, then applied Bonferroni corrections to account for multiple comparisons. To compare means, we used z-tests. We conducted statistical analyses using the R programming environment, version 4.2.3 (R Foundation for Statistical Computing, Vienna, Austria).

## RESULTS

### Participant Characteristics

During the review period, we identified 491,048 women with a diagnosis of endometriosis across the CCAE (N=236,594), MDCD (N=70,426), Optum® EHR (N=181,743), and CUIMC EHR (N=2,285) datasets (**Figure S1**). Across the four databases, participants ranged in age from 15-19 years old in 2-3% of cases, 20-24 years in 5-9%, 25-29 years in 10-14%, 30-34 years in 15-19%, 35-39 years in 20-22%, 40-44 years in 20-25%, and 45-49 years old in 14-23% of cases (**Table 1, Tables S1-S3**). Women were identified as 1% Asian, 7-8% Black or African American, and 37-40% white depending on the data source; 52-55% were of unknown race and 3% were of Hispanic or Latina ethnicity. The CCAE dataset did not provide race or ethnicity data.

**Table 1.** Participant Characteristics, Optum EHR Dataset.

| Characteristic | Cohort A<br>(N = 100,697) | Cohort B<br>(N=48,241) | Cohort C<br>(N = 128,879) | Cohort D<br>(N = 133,819) | Cohort E<br>(N = 139,368) |
| --- | --- | --- | --- | --- | --- |
| **Age Group** |  |  |  |  |  |
| 15-19 years | 1,290 (1.3%) | 1,169 (2.4%) | 2,827 (2.2%) | 2,937 (2.2%) | 3,023 (2.2%) |
| 20-24 years | 5,051 (5.0%) | 4,242 (8.8%) | 9,568 (7.4%) | 10,024 (7.5%) | 10,362 (7.4%) |
| 25-29 years | 9,785 (9.7%) | 7,239 (15%) | 16,600 (13%) | 17,397 (13%) | 18,028 (13%) |
| 30-34 years | 15,720 (16%) | 9,504 (20%) | 23,381 (18%) | 24,276 (18%) | 25,120 (18%) |
| 35-39 years | 20,619 (20%) | 10,006 (21%) | 27,294 (21%) | 28,391 (21%) | 29,546 (21%) |
| 40-44 years | 24,973 (25%) | 9,376 (19%) | 27,549 (21%) | 28,459 (21%) | 29,784 (21%) |
| 45-49 years | 23,259 (23%) | 6,705 (14%) | 21,660 (17%) | 22,335 (17%) | 23,505 (17%) |
| **Race** |  |  |  |  |  |
| Asian | 1,976 (2.0%) | 866 (1.8%) | 2,155 (1.7%) | 2,246 (1.7%) | 2,370 (1.7%) |
| Black / African American | 11,231 (11%) | 5,726 (12%) | 14,875 (12%) | 15,308 (11%) | 15,866 (11%) |
| White | 81,420 (81%) | 38,597 (80%) | 103,353 (80%) | 107,365 (80%) | 111,729 (80%) |
| Unknown | 6,070 (6.0%) | 3,052 (6.3%) | 8,496 (6.6%) | 8,900 (6.7%) | 9,403 (6.7%) |
| **Ethnicity** |  |  |  |  |  |
| Hispanic / Latina | 6,460 (6.4%) | 3,403 (7.1%) | 8,520 (6.6%) | 8,810 (6.6%) | 9,114 (6.5%) |
| Not Hispanic / Latina | 87,584 (87%) | 41,713 (86%) | 111,929 (87%) | 116,076 (87%) | 120,767 (87%) |
| Unknown | 6,653 (6.6%) | 3,125 (6.5%) | 8,430 (6.5%) | 8,933 (6.7%) | 9,487 (6.8%) |

### Cohort Characteristics

In total, phenotyping approaches identified 278,971 women with endometriosis in Cohort A, 157,959 in Cohort B, 363,049 in Cohort C, 373,201 in Cohort D, and 377,036 in Cohort E (**Figure S1**). A sizable proportion of women (15-20% of endometriosis cases) were identified across all 5 cohorts regardless of phenotype definition (**Figure 1**, **Table S4**). More than one-fourth (26-30%) were represented in symptom-only Cohorts C-E but not in Cohorts A and B (**Table S4**). We also note that 2-4.6% of cases (14,795 women total) showed only pelvic and/or abdominal pain but not other symptoms noted in clinical guidelines (**Table S4**). Largely, our findings are consistent across each dataset. In the interest of brevity, remaining findings presented in the main text are from our largest EHR dataset (Optum® EHR). Additional tables and figures, including those based on data from other sources, are provided as Supplemental Materials (**Tables S1-S11**, **Figures S2-S22**).

**Figure 1a-d.**
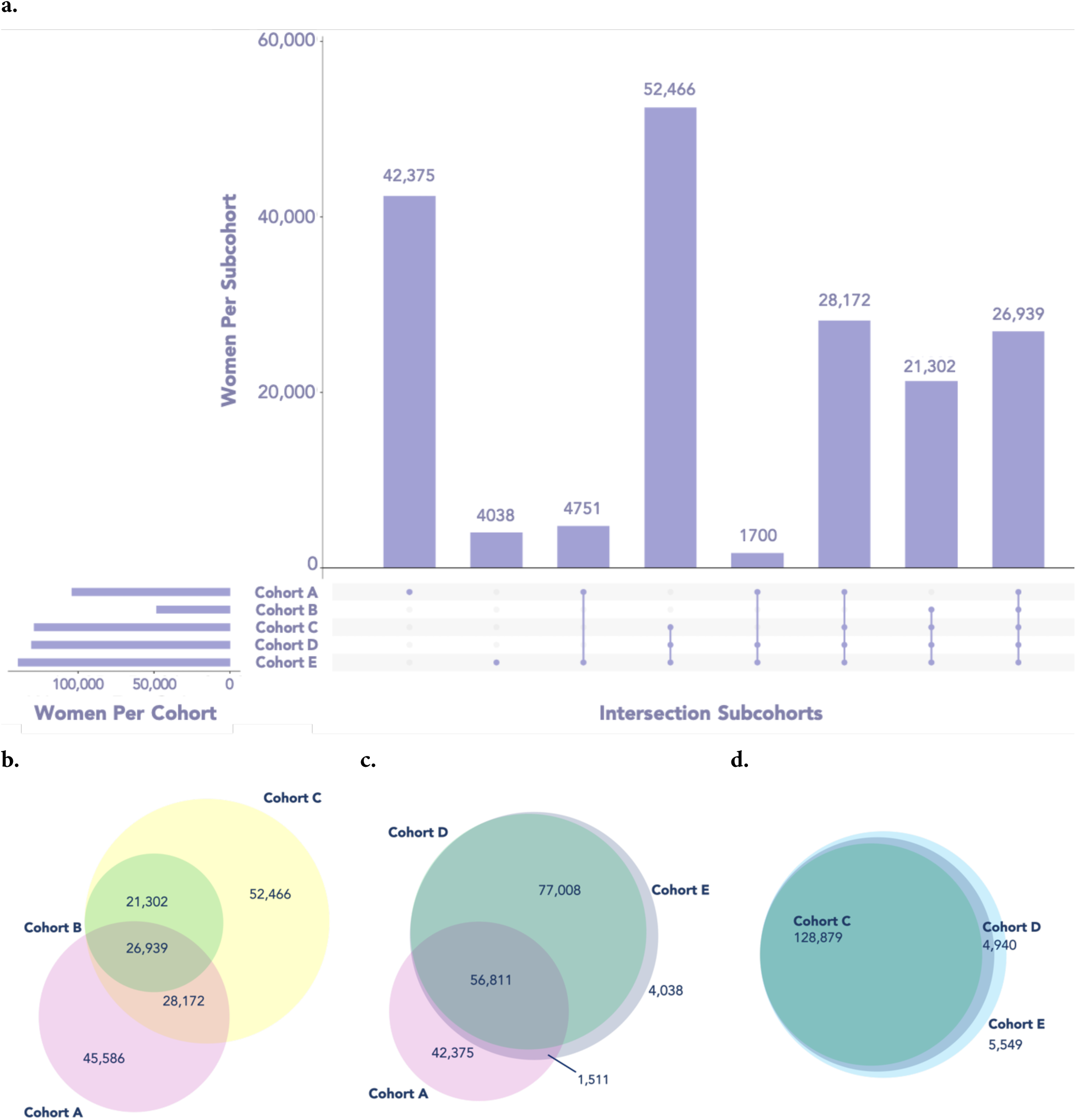
Patient Set Overlap Among Study Cohorts in Optum EHR Dataset. a) UpSet plot illustrating intersections among patients identified across all cohorts. b) Venn diagram of patients in Cohorts A, B, and C. c) Venn diagram of patients in Cohorts A, D, and E. d) Venn diagram of patients in Cohorts C, D, and E. Cohort A: Surgical confirmation phenotype; Cohort B: Imaging and guideline-recognized symptoms phenotype; Cohort C: Guideline-recognized symptoms only phenotype; Cohort D: Guideline-recognized symptoms and/or pelvic pain phenotype; Cohort E: Guideline-recognized symptoms, pelvic pain, and/or abdominal pain phenotype. *Abbreviations:* EHR, Electronic Health Record.

### Timing of Diagnosis by Cohort

#### Age at Diagnosis

Women with a diagnosis of endometriosis based on surgical confirmation (Cohort A) were older at time of diagnosis (mean age = 38 years, SD = 8) compared to women diagnosed based on imaging and symptoms (Cohort B mean age = 35, SD = 9; p<0.001, **Figure 2b**). We found a similar trend when comparing age at diagnosis between Cohort A and Cohorts C-E (mean age = 36, average SD = 8) with a mean age difference of 2 years (p<0.001 for all comparisons). This pattern was consistent across all four data sources (**Figure 2**, **Figures S5-S7**).

**Figure 2a-d.**
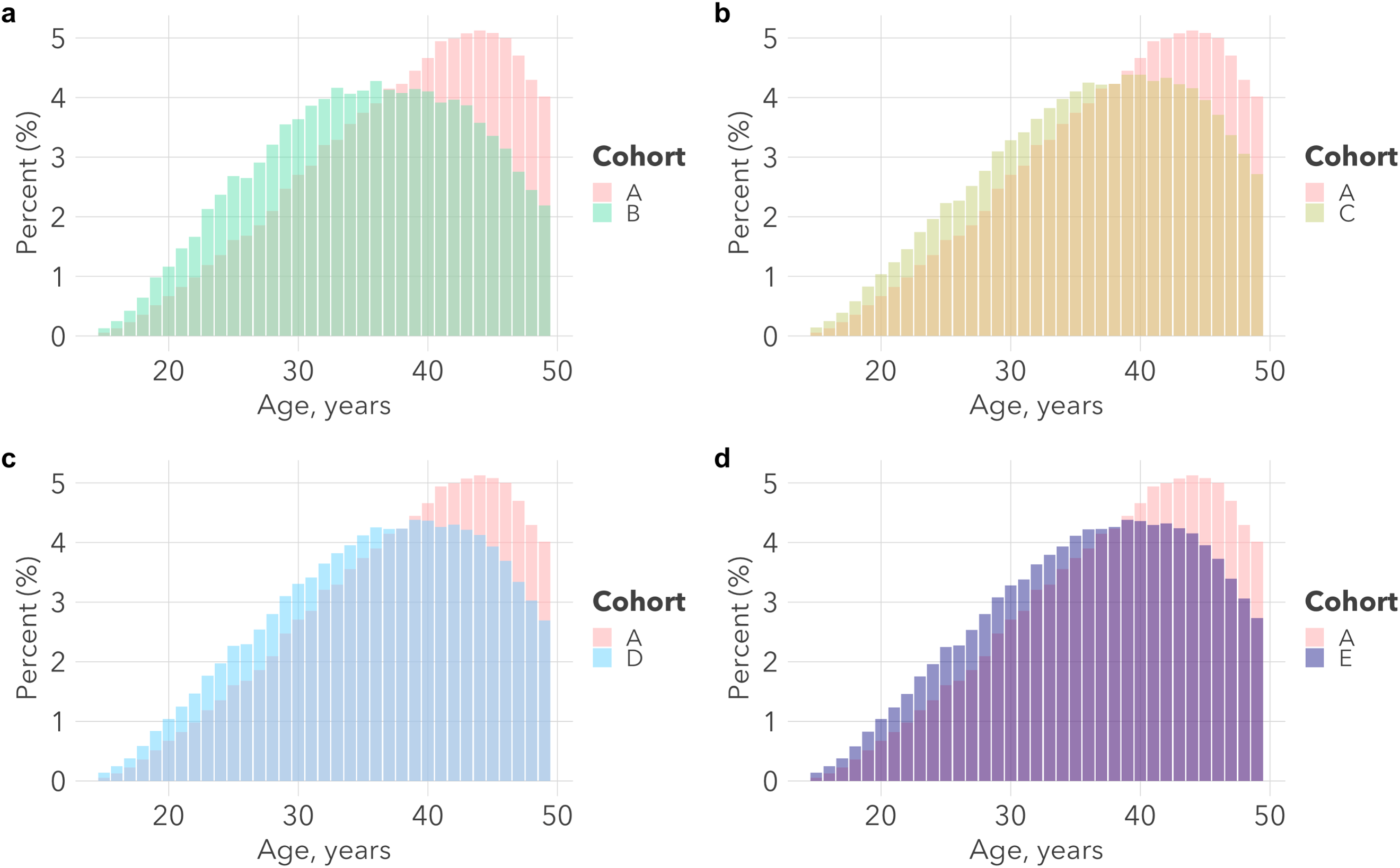
Age at Diagnosis of Endometriosis in the Optum EHR Dataset. On average, women in Cohort A were diagnosed with endometriosis at an older age than women in Cohorts B-E (mean age difference Cohort A vs Cohorts C-E = 2 years; p<0.001 for all comparisons). a) Age at diagnosis among patients in Cohorts A and B. b) Age at diagnosis among patients in Cohorts A and C. c) Age at diagnosis among patients in Cohorts A and D. d) Age at diagnosis among patients in Cohorts A and E. Cohort A: Surgical confirmation phenotype; Cohort B: Imaging and guideline-recognized symptoms phenotype; Cohort C: Guideline-recognized symptoms only phenotype; Cohort D: Guideline-recognized symptoms and/or pelvic pain phenotype; Cohort E: Guideline-recognized symptoms, pelvic pain, and/or abdominal pain phenotype. *Abbreviations:* EHR, Electronic Health Record.

#### Delay in Diagnosis

Among women identified with endometriosis, we also compared their date of entry into Cohort A with date of entry into Cohorts B-E (**Figure 3**, **Figures S8-S10**). For most women, there was little to no difference in date of cohort entry between Cohort A and Cohort B (median difference = 0 days; IQR = 0-45 days). Similarly, we found almost no difference between entry into Cohort A and Cohorts C-E (median difference = 0 days for all, IQR = 0-15 days for Cohort C, 0-14 days for Cohort D, and 0-13 days for Cohort E).

**Figure 3a-d.**
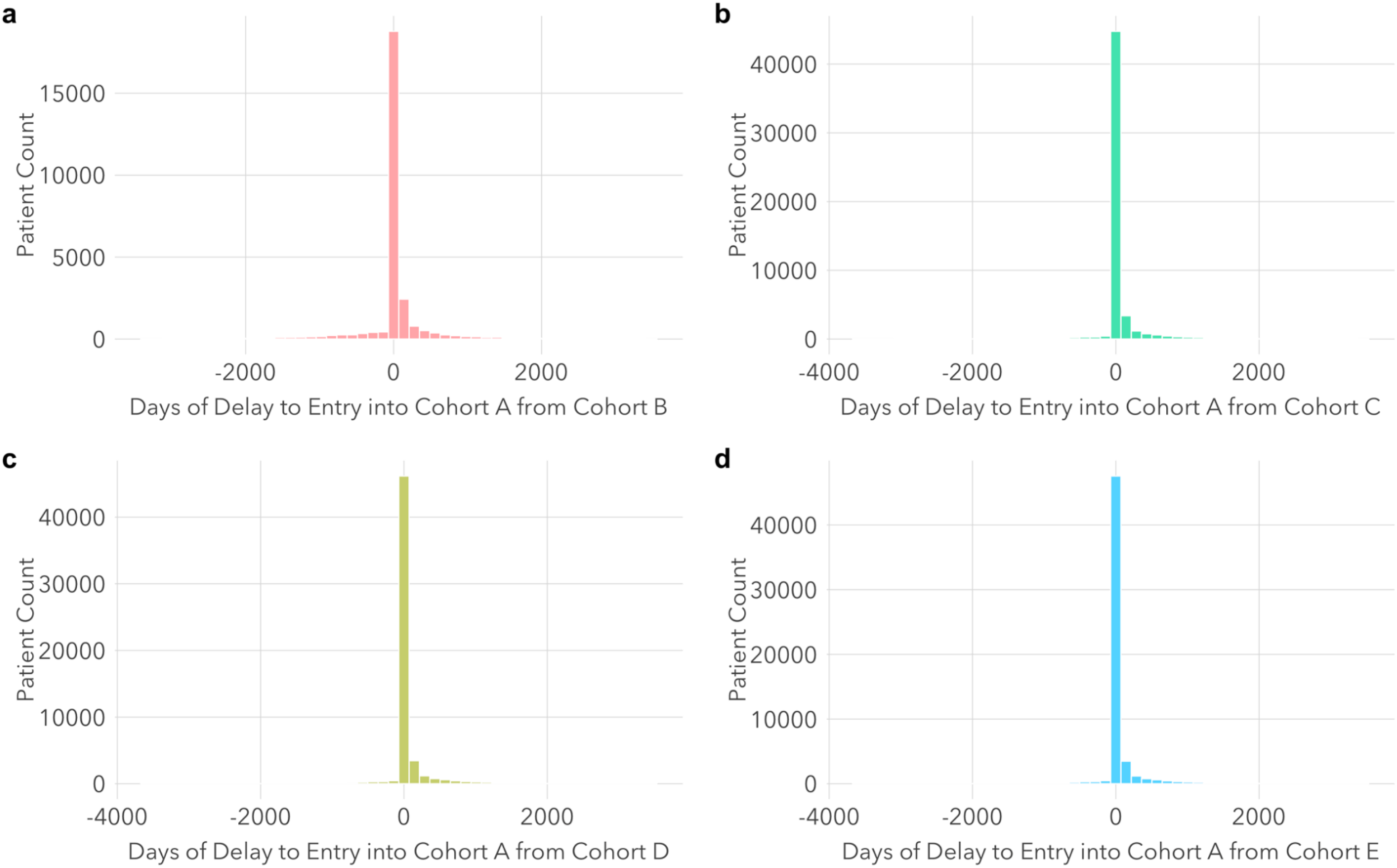
Differences in Cohort Entry Dates in the Optum EHR Dataset. Among most women identified by more than one phenotype definition, there was little to no difference in date of cohort entry (e.g., median difference in entry date between Cohort A and B = 0 days, interquartile range [IQR] = 0-45 days). a) Differences in cohort entry dates among patients identified by both cohort definitions A and B. b) Differences in cohort entry dates among patients identified by both cohort definitions A and C. c) Differences in cohort entry dates among patients identified by both cohort definitions A and D. d) Differences in cohort entry dates among patients identified by cohort definitions A and E. Cohort A: Surgical confirmation phenotype; Cohort B: Imaging and guideline-recognized symptoms phenotype; Cohort C: Guideline-recognized symptoms only phenotype; Cohort D: Guideline-recognized symptoms and/or pelvic pain phenotype; Cohort E: Guideline-recognized symptoms, pelvic pain, and/or abdominal pain phenotype. *Abbreviations:* EHR, Electronic Health Record.

### Clinical Histories by Cohort

#### Conditions and Symptoms

The top 25 conditions with the greatest prevalence difference between Cohort A and Cohort B and between Cohort A and Cohort E are shown in **Figure 4a-b** (see **Figures S11-S14** for comparisons between Cohort A and Cohorts C and D). The most common symptom experienced by women prior to receipt of an endometriosis diagnosis was pain. Pain was reported among 91% of women in Cohort A and 100% of women in Cohorts B-E. Among all five cohorts, localized pain was most frequently reported as abdominal pain and pain in the pelvis with prevalence ranging from 69% to 70% of women in Cohort A compared to 83% to 90% in Cohorts B-E.

**Figure 4a-b.**
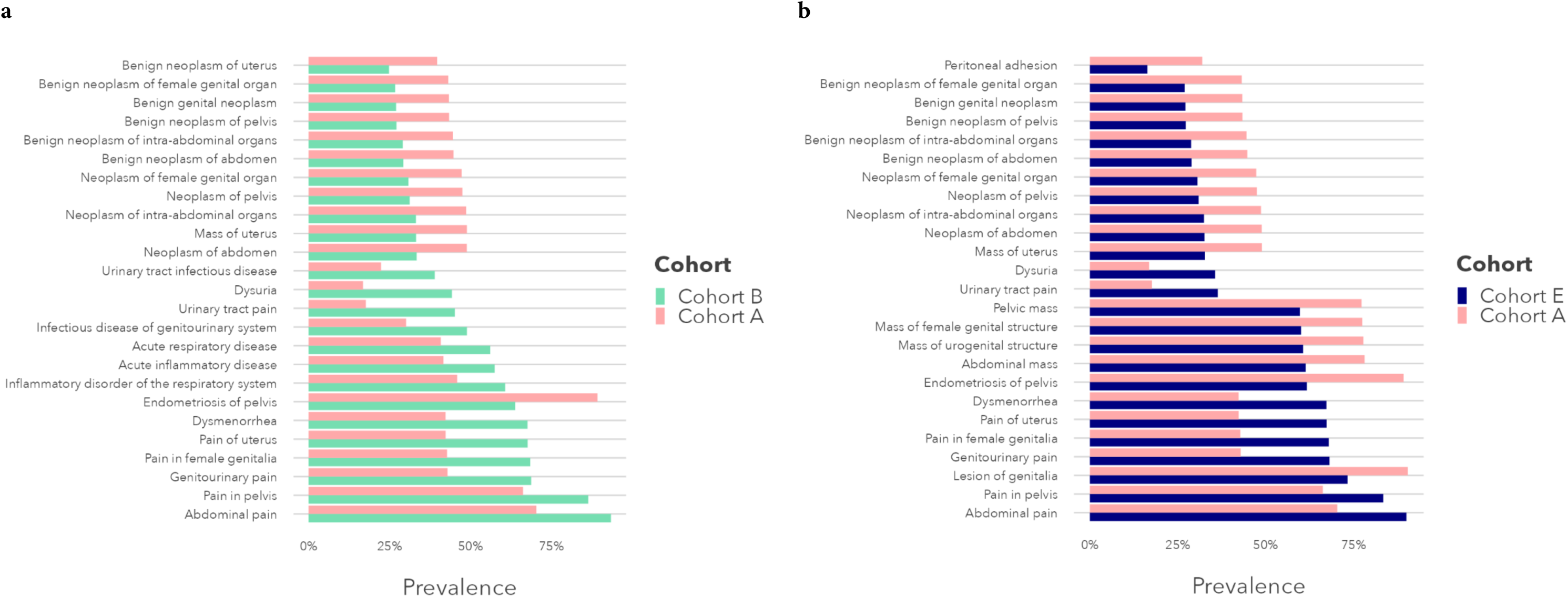
Condition Concepts in the Optum EHR Dataset with the Largest Differences in Prevalence Between Endometriosis Cohorts Based on Surgical Diagnosis (Cohort A) and Imaging and/or Symptoms-based Diagnosis (Cohorts B and E). Compared to Cohort A, localized pain (e.g., abdominal, pelvic, genitourinary), dysmenorrhea, and dysuria were consistently more common in Cohorts B and E. Lesions, masses, and neoplasms (e.g., abdominal, pelvic, uterine) were more common in Cohort A. Cohort A: Surgical confirmation phenotype; Cohort B: Imaging and guideline-recognized symptoms phenotype; Cohort E: Guideline-recognized symptoms, pelvic pain, and/or abdominal pain phenotype. *Abbreviation:*EHR: electronic health record.

Genitourinary pain (43% vs 68-69%), pain in female genitalia (43% vs 67-68%), and pain of uterus (42% vs 67-68%) were also more commonly reported among patients identified based on imaging and/or symptom presentation (Cohorts B-E). We also found that urinary tract pain was documented more than twice as much among women in Cohorts B-E (36-45% of women) as Cohort A (18%). Condition codes for infectious disease of genitourinary system (30% in Cohort A, 49% in Cohort B, 39-41% in Cohorts C-E) and urinary tract infectious disease (22% in Cohort A, 39% in Cohort B, 31-32% in Cohorts C-E) were also more frequently reported in cohorts based on imaging and/or symptoms.

Similarly, we found substantial documentation of dysuria (17% in Cohort A, 37%-44% in Cohorts B through E) and dysmenorrhea (42% of patients in Cohort A, 67%-68% in Cohorts B-E) in these cohorts. Dyspareunia was recorded in nearly one-sixth to one-fourth of cases (16% in Cohort A, 24-27% in Cohorts B-E) and fatigue in one-sixth to one-fifth of cases (15% in Cohort A, 18-22% in Cohorts B-E). The non-specific diagnosis of acute inflammatory disease was also more common among these cohorts (42% vs 49-57%) as were conditions not traditionally associated with endometriosis such as inflammatory disorder of the respiratory system (46% vs 53-61%) and acute respiratory disease (41% vs 48-56%).

In contrast, we found more frequent documentation of lesions, masses, and neoplasms in Cohort A compared to Cohorts B-E. For example, lesions of the genitalia (prevalence range: 73-90%), ovary (46-59%), and uterus (50-68%), abdominal mass (61-78%), mass of urogenital structure (61-78%), pelvic mass (60-77%), and mass of uterus (33-49%) were more commonly identified among women diagnosed by surgical confirmation (Cohort A) than the other phenotype definitions (Cohorts B-E). We also noted significant differences between neoplasms of the abdomen (33-49%), female genital organs (31-47%), intra-abdominal organs (32-49%), uterus (29-44%), and pelvis (31-48%). Reporting of peritoneal adhesions was twice as common in Cohort A (32%) compared to Cohorts B-E (16-18%).

#### Medications

The top 25 medications with the greatest prevalence difference between Cohort A and Cohorts B through E are shown in **Figure 5** and **Figures S15-S18.** Among treatments recommended by the ESHRE guidelines for endometriosis-related symptoms, women in Cohorts B-E were significantly more likely to receive hormones and related agents (44% in Cohort A, 61% in Cohort B, 50-51% in Cohorts C-E), including hormonal contraceptives for systemic use (50% in Cohort A, 67% in Cohort B, 56-57% in Cohorts C-E), compared to women in Cohort A. We also found that opioids had high levels of utilization among all cohorts but were most common among women in Cohort A (96% in Cohort A, 82% in Cohort B, 77-78% in Cohorts C-E). The use of antiemetics and antinauseants (71-93%), antipruritics (68-88%), antacids (62-89%), and corticosteroids (70-87%) were also high overall and highest in Cohort A. Antidiarrheal microorganisms (67-90%), other antidiarrheals (57-87%), and urologicals (62-88%) were also most prevalent among women in Cohort A.

**Figure 5a-b.**
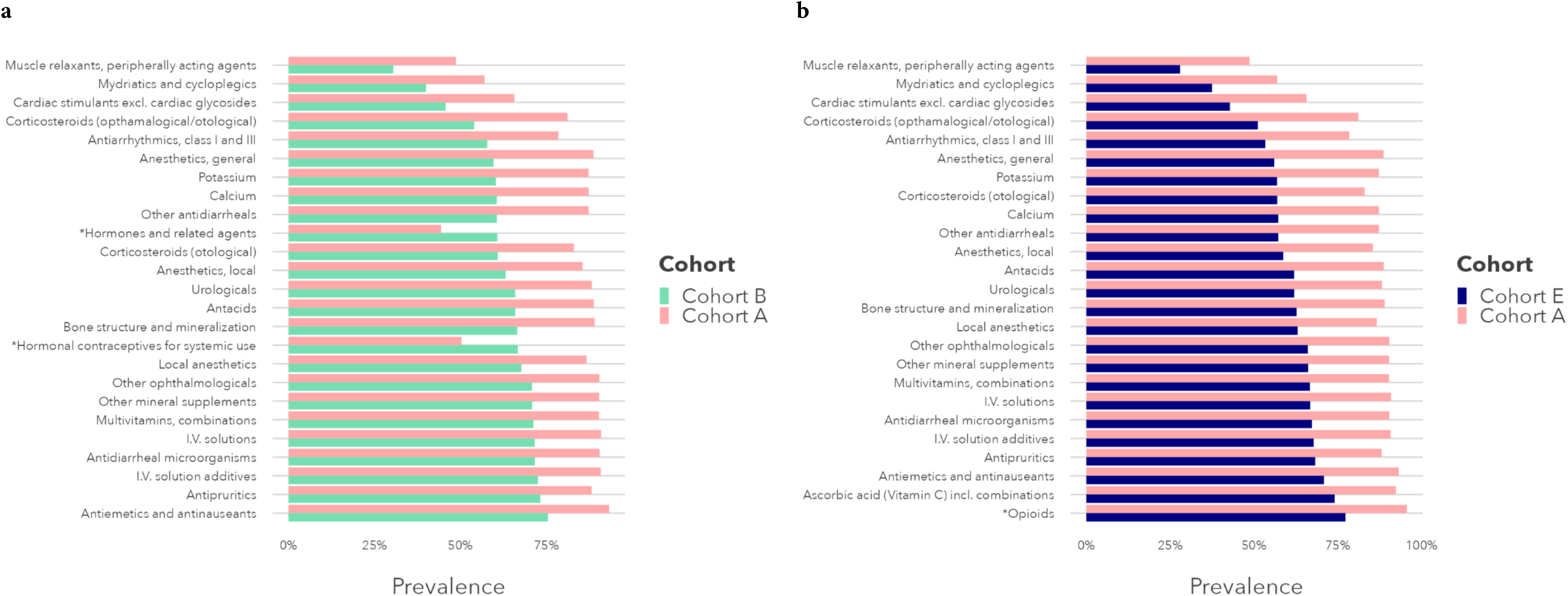
Medication Concepts in the Optum EHR Dataset with the Largest Differences in Prevalence Between Endometriosis Cohorts Based on Surgical Diagnosis (Cohort A) and Imaging and/or Symptoms-based Diagnosis (Cohorts B and E). Nonsteroidal anti-inflammatory and antirheumatics, other analgesics, corticosteroids, and drugs for gastrointestinal disorders were more common in Cohort A. Hormones, antidepressants, anxiolytics, and antibacterials were more common in Cohorts B and E. *Denotes medications included among ESHRE treatment endometriosis guidelines. Cohort A: Surgical confirmation phenotype; Cohort B: Imaging and guideline-recognized symptoms phenotype; Cohort E: Guideline-recognized symptoms, pelvic pain, and/or abdominal pain phenotype. *Abbreviation:* EHR: electronic health record.

#### Procedures

Notably, routine gynecologic examination was only recorded in slightly more than one-third of patients (34% in Cohort A, 44% in Cohort B, and 35-36% in Cohorts C-E). The top procedures with the greatest prevalence difference between cohorts are illustrated in **Figure 6** and **Figures S19-S22.** We found that women with a diagnosis of endometriosis based on surgical confirmation (Cohort A) were more likely to have received laparoscopy with fulguration or excision of lesions of the lower abdomen (38% vs 15-21%), laparoscopy with total hysterectomy for uterus 250g or less (30% vs 7-9%), and surgical hysteroscopy with biopsy of endometrium (12% vs 7-8%). We also note that surgical pathology with microscopic evaluation was performed more than twice as often in Cohort A (48% in Cohort A, 16% in Cohort B, and 21% in Cohorts C-E).

**Figure 6a-b.**
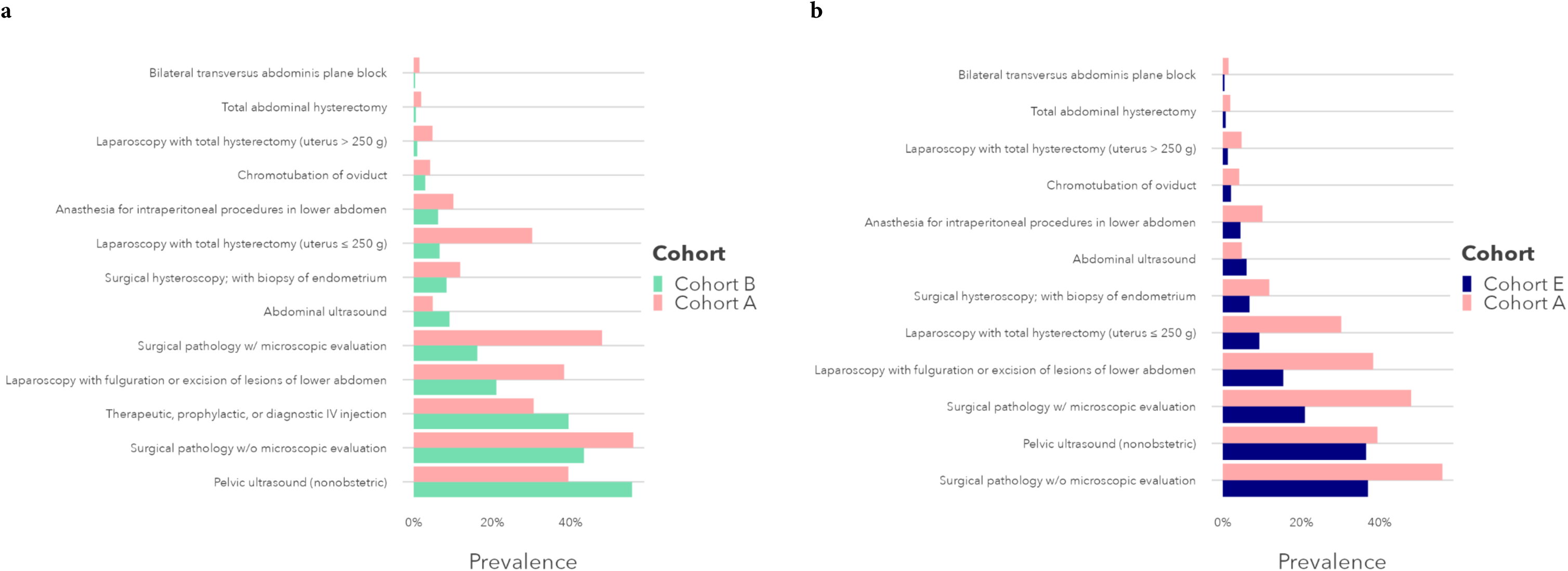
Procedure Concepts in the Optum EHR Dataset with the Largest Differences in Prevalence Between Endometriosis Cohorts Based on Surgical Diagnosis (Cohort A) and Imaging and/or Symptoms-based Diagnosis (Cohorts B and E). Among differences found, women with a diagnosis of endometriosis based on surgical confirmation (Cohort A) were more likely to have received laparoscopy with total hysterectomy or surgical pathology with microscopic evaluation. Cohort A: Surgical confirmation phenotype; Cohort B: Imaging and guideline-recognized symptoms phenotype; Cohort E: Guideline-recognized symptoms, pelvic pain, and/or abdominal pain phenotype. *Abbreviation:* EHR: electronic health record.

Transvaginal ultrasound was performed the most among procedures conducted, especially among patients in Cohort B (60% in Cohort A, 84% in Cohort B, and 53-55% in Cohorts C-E). Non-obstetric pelvic ultrasound was conducted for slightly more than one-half of patients in Cohort B but only roughly one-third in other cohorts (39% in Cohort A, 56% in Cohort B, and 37% in Cohorts C-E). We found that abdominal ultrasound was performed infrequently though most often among patients in Cohort B (5% in Cohort A, 9% in Cohort B, 6% in Cohorts C-E). Therapeutic, prophylactic, or diagnostic intravenous injection was also more common in Cohort B than all other cohorts (40% vs 31-32%).

#### Healthcare Utilization

Before receiving an endometriosis diagnosis, nearly all participants in each cohort had at least one outpatient visit (**Tables S8-S11**). Compared to women diagnosed with endometriosis based on surgical confirmation, more women diagnosed based on imaging and symptom presentation had emergency room (ER) visits (41% of women in Cohort B vs 30% in Cohort A, p<0.001). They also used the ER with greater frequency prior to diagnosis (mean number of ER visits = 1.4, SD = 4.3 in Cohort B vs mean of 0.8 ER visits, SD=3.2 in Cohort A; p<0.001). While nearly half of women (45%) in Cohort A had at least one hospitalization prior to diagnosis (mean of 0.6 visits, SD=1.1), this was the case for only one-fourth (26%) in Cohort B (mean of 0.4 visits, SD=1.8). Similar trends held when comparing Cohorts C-E to Cohort A and across all datasets. Notably, women in the MDCD dataset had consistently higher levels of healthcare utilization overall.

## DISCUSSION

Our study of four large national insurance claims and EHR databases characterized differences in clinical histories and demographics for nearly half a million women diagnosed with endometriosis in the US. Findings from our analysis suggest that the latest shift in clinical guidelines results in more comprehensive diagnosis of women with the disease, but updated ESHRE diagnostic criteria still do not capture a sizable percentage of women with endometriosis. Most notably, despite using phenotypes with high levels of precision (PPV), only 15-20% of all endometriosis cases were shared among all five cohorts. This illustrates both the heterogeneous presentation of the disease as well as the importance of expanding the criteria used for diagnosis.

Our analysis focuses on comparison of cohorts based on five complementary phenotype definitions, two of which operationalize the original and updated diagnostic criteria delineated in the ESHRE clinical guidelines (Cohorts A and B, respectively). Importantly, the most recent ESHRE guideline asserts that, while imaging via ultrasound or MRI are recommended in a diagnostic work-up, a negative finding does not necessarily rule out endometriosis as a potential diagnosis.^9,14–18^ In light of this, we took a data-driven approach that identified a substantial cohort of patients whose literal pain would have been ignored by either cohort definition. This manifested as three additional symptom-only cohorts based on phenotype definitions that first built upon the current guideline by incorporating only ESHRE-recognized symptoms (Cohort C) with successive expansion to include either one or both of the two most commonly reported symptoms of endometriosis – pelvic pain and/or abdominal pain (Cohorts D and E, respectively).

Importantly, cohorts derived from the newer guidelines and its expanded iterations identified patients who were younger at the time of diagnosis. Given that the disease is suspected to be congenital and prior studies suggest a diagnostic delay of four to eleven years from first symptom onset, this finding is not inconsiderable. While the new guidelines were only released a short time ago, this also nonetheless illustrates substantial room for improvement. Further demonstrating this is our comparison of differences in the timing of cohort entry. We found little-to-no difference in entry date between symptom cohorts, with some indication that entry to the surgical cohort skewed slightly toward later dates, representing a diagnostic delay. Additionally, the racial and ethnic makeups of patients identified under the new guidelines were largely unchanged, though notably women who identified as African American or Black routinely represented a quarter of patients in each cohort segment in the Medicaid dataset while only comprising 15% of the US population of women.

In light of its heterogenous presentation, flexible guidelines are especially necessary to prevent under– and delayed diagnosis and any potential disparities that may have arisen in the past by maintaining surgical confirmation as a strict requirement.^13^ Given the complexity of endometriosis, it is also paramount that as additional evidence is generated, novel insights are further incorporated into clinical practice. This transformation also has important implications for how observational health research is conducted. Changes in clinical documentation will require updating existing approaches to identify cohorts of patients with endometriosis and those cohort definitions, if not rigorously examined, may in turn result in skewed characterizations of women with the disease.

We found substantial differences in the prevalence of conditions, medications, and procedures associated with patients in each cohort. Primarily these significant differences were between Cohort A and all the other cohorts. Nonetheless, despite the similarities in composition of Cohorts B-E, there were still many women not represented under the new guidelines. Our investigation found that nearly one-fourth of cases were consistently identified only when using the symptom-based phenotypes (Cohorts C-E), suggesting that imposing surgery or imaging-related constraints to diagnosis may miss a substantial portion of women with endometriosis. Further, a small but sizable percentage of women (nearly 15,000 cases) presented with only pelvic and/or abdominal pain and none of the other guideline-recognized symptoms.

Among conditions for which there was significant difference between Cohort A and Cohorts B-E, we noted that urinary tract pain was both commonly documented among women in Cohorts B-E and at rates more than twice that of women in Cohort A. Additionally, condition codes for infectious disease of genitourinary system and urinary tract infectious disease were much more frequently reported in Cohorts B-E. While we cannot be certain based on available data, one potential hypothesis is that when women present with urogenital pain, in the absence of surgical, imaging, or laboratory test confirmation, clinicians may first suspect urinary tract infection instead of endometriosis, resulting in diagnostic delay.

Our analysis also revealed differences in patterns of healthcare utilization among women diagnosed with endometriosis. For example, we observed higher rates of ER visits among women in cohorts based on imaging and/or symptom presentation compared to surgical confirmation. These findings underscore the continued need for improved access to timely and appropriate care for women with endometriosis, particularly those with non-classical symptoms, different care-seeking preferences, or those lacking access to surgical intervention.

### Limitations

While our study provides valuable insights regarding the clinical characteristics of endometriosis among women in the US, we recognize several limitations that should be acknowledged. First, our retrospective observational study design inherently limits our ability to establish causality. As secondary data sources, insurance claims and EHR data also introduce potential biases and inaccuracies inherent to administrative data coding and documentation practices. Second, while we largely evaluate differences among cohorts within each data source separately to preserve internal consistency, there is non-zero possibility that a woman represented in one data source may also be represented in another and we do not account for potential correlation. Third, while we did not detect any statistically significant differences in race or ethnicity between cohorts, there were a large percentage of women of unknown race or ethnicity. Fourth, our phenotyping algorithms rely on condition, observation, and procedure codes that did not necessarily capture the full spectrum of diagnostic criteria noted in the ESHRE guidelines (i.e., dyschezia, cyclical cough, and cyclical scar swelling and pain). Further, algorithmic performance may also differ across healthcare settings and populations. Fifth, by design, our inclusion criteria for each cohort were based on specific combinations of diagnostic criteria and may limit the generalizability of our findings to all women with endometriosis, particularly among those with atypical presentations. Lastly, each of our phenotypes required diagnosis codes for endometriosis. Our approach likely underestimates the true prevalence of endometriosis, as the condition is known to be underdiagnosed and underreported in clinical practice.

### Conclusion

Our study provides novel insights into the complex interplay between diagnostic criteria, clinical characteristics, and healthcare utilization patterns among women diagnosed with endometriosis in the US. By elucidating these relationships, our findings contribute to a deeper understanding of the heterogeneity of endometriosis and underscore the importance of personalized approaches to diagnosis and management. Moving forward, efforts to optimize diagnostic algorithms, expand symptom recognition, and enhance access to comprehensive care are critical to addressing the unmet needs of women affected by this condition.

### Data Availability Statement

Merative™ MarketScan® claims databases and Optum® de-identified EHR data set (Optum® EHR) are publicly available for research purposes but require a license and access provided by a third party. The electronic health record and administrative data from Columbia University Irving Medical Center are not available due to restrictions to preserve patient confidentiality.

### Authors’ Roles

All authors had full access to the data in the study. They also take responsibility for the integrity of the data and accuracy of the analysis and have approved the final manuscript.

Concept and design: Elhadad, Reyes Nieva, Kashyap, Ryan.

Acquisition, analysis, or interpretation of data: All authors Drafting of the manuscript: Reyes Nieva, Elhadad.

Critical revision of the manuscript for important intellectual content: All authors

Statistical analysis: Reyes Nieva, Kashyap, Anand, Ketenci.

Obtained funding: Elhadad.

Administrative, technical, or material support: Reyes Nieva, Voss, Kashyap, Anand, Ketenci, Ostropolets, Choi, DeFalco, Li, Allen, Guang.

Supervision: Elhadad, Ryan.

### Funding

The research was supported in part by grants from the National Library of Medicine and National Heart, Lung, and Blood Institute at the National Institutes of Health (T15-LM007079 [HRN, AK, AA]; R01HL148248 [MK]; R01LM013043 [NE]) and a Computational and Data Science Fellowship from the Association for Computing Machinery Special Interest Group in High Performance Computing (HRN). Funding sources had no role in study design; in the collection, analysis and interpretation of data; in the writing of the report; or in the decision to submit the article for publication.

## Supplementary Methods

**Supplementary Results**

**Supplementary Discussion**

### Supplementary Tables

● Table S1. Participant Characteristics, Merative Commercial Claims and Encounters (CCAE) Dataset
● Table S2. Participant Characteristics, Merative Medicaid (MDCD) Dataset
● Table S3. Participant Characteristics, Columbia University Irving Medical Center (CUIMC) EHR Dataset
● Table S4. Patient Set Overlap Among Study Cohorts, All Datasets
● Table S5. Patient Race and Ethnicity Among Cohort Intersections, Optum EHR Dataset
● Table S6. Patient Race and Ethnicity Among Cohort Intersections, MDCD Dataset
● Table S7. Patient Race and Ethnicity Among Cohort Intersections, CUIMC EHR Dataset
● Table S8. Healthcare Utilization, Optum EHR Dataset
● Table S9. Healthcare Utilization, CCAE Dataset
● Table S10. Healthcare Utilization, MDCD Dataset
● Table S11. Healthcare Utilization, CUIMC EHR Dataset
● Table S12. Classification Report for Endometriosis Cohort Definitions
● Table S13. Pairwise Percent Agreement Between Cohorts in Chart Review Sample
● Table S14. Pairwise Cohen’s Kappa Scores Between Cohorts in Chart Review Sample

### Supplementary Figures

● Figure S1. STROBE Flow Diagram of Women Diagnosed with Endometriosis Based on Surgery, Imaging, and/or Symptom Presentation in the CCAE, Optum EHR, MDCD, and CUIMC EHR Databases
● Figure S2a-d. Patient Set Overlap Among Study Cohorts in CCAE Dataset
● Figure S3a-d. Patient Set Overlap Among Study Cohorts in MDCD Dataset
● Figure S4a-d. Patient Set Overlap Among Study Cohorts in CUIMC Dataset
● Figure S5a-d. Age at Diagnosis of Endometriosis in the CCAE Dataset
● Figure S6a-d. Age at Diagnosis of Endometriosis in the MDCD Dataset
● Figure S7a-d. Age at Diagnosis of Endometriosis in the CUIMC Dataset
● Figure S8a-d. Differences in Cohort Entry Dates in the CCAE Dataset
● Figure S9a-d. Differences in Cohort Entry Dates in the MDCD Dataset
● Figure S10a-d. Differences in Cohort Entry Dates in the CUIMC Dataset
● Figure S11a-b. Condition Concepts in the Optum EHR Dataset with the Largest Differences in Prevalence between Endometriosis Cohorts Based on Surgical Diagnosis (Cohort A) and Symptoms-based Diagnosis (Cohorts C and D)
● Figure S12a-d. Condition Concepts in the CCAE Dataset with the Largest Differences in Prevalence between Endometriosis Cohorts Based on Surgical Diagnosis (Cohort A) and Imaging and/or Symptoms-based Diagnosis (Cohorts B-E)
● Figure S13a-d. Condition Concepts in the MDCD Dataset with the Largest Differences in Prevalence between Endometriosis Cohorts Based on Surgical Diagnosis (Cohort A) and Imaging and/or Symptoms-based Diagnosis (Cohorts B-E)
● Figure S14a-d. Condition Concepts in the CUIMC EHR Dataset with the Largest Differences in Prevalence between Endometriosis Cohorts Based on Surgical Diagnosis (Cohort A) and Imaging and/or Symptoms-based Diagnosis (Cohorts B-E)
● Figure S15a-b. Medication Concepts in the Optum EHR Dataset with the Largest Differences in Prevalence between Endometriosis Cohorts Based on Surgical Diagnosis (Cohort A) and Symptoms-based Diagnosis (Cohorts C and D)
● Figure S16a-d. Medication Concepts in the CCAE Dataset with the Largest Differences in Prevalence between Endometriosis Cohorts Based on Surgical Diagnosis (Cohort A) and Imaging and/or Symptoms-based Diagnosis (Cohorts B-E)
● Figure S17a-d. Medication Concepts in the MDCD Dataset with the Largest Differences in Prevalence between Endometriosis Cohorts Based on Surgical Diagnosis (Cohort A) and Imaging and/or Symptoms-based Diagnosis (Cohorts B-E)
● Figure S18a-d. Medication Concepts in the CUIMC EHR Dataset with the Largest Differences in Prevalence between Endometriosis Cohorts Based on Surgical Diagnosis (Cohort A) and Imaging and/or Symptoms-based Diagnosis (Cohorts B-E)
● Figure S19a-b. Procedure Concepts in the Optum EHR Dataset with the Largest Differences in Prevalence between Endometriosis Cohorts Based on Surgical Diagnosis (Cohort A) and Symptoms-based Diagnosis (Cohorts C and D)
● Figure S20a-d. Procedure Concepts in the CCAE Dataset with the Largest Differences in Prevalence between Endometriosis Cohorts Based on Surgical Diagnosis (Cohort A) and Imaging and/or Symptoms-based Diagnosis (Cohorts B-E)
● Figure S21a-d. Procedure Concepts in the MDCD Dataset with the Largest Differences in Prevalence between Endometriosis Cohorts Based on Surgical Diagnosis (Cohort A) and Imaging and/or Symptoms-based Diagnosis (Cohorts B-E)
● Figure S22a-d. Procedure Concepts in the CUIMC EHR Dataset with the Largest Differences in Prevalence between Endometriosis Cohorts Based on Surgical Diagnosis (Cohort A) and Imaging and/or Symptoms-based Diagnosis (Cohorts B-E)

### Supplementary Methods

We conducted a detailed chart review to validate and compare the relative performance of our five endometriosis cohort definitions. As it afforded us the ability to examine the full electronic health record (EHR) for patient care received at Columbia University Irving Medical Center (CUIMC), we generated cohorts for each phenotype definition from the CUIMC EHR dataset and selected a stratified random sample of 100 patients satisfying criteria for our cohort definitions. We selected this sample size using established best practices for EHR-based phenotype algorithm development.^19,20^ Due to the de-identified nature and our lack of access to their originating data sources, we were unable to perform equivalent chart review on the Merative™ MarketScan® claims and Optum® de-identified EHR (Optum® EHR) databases.

To establish cases and controls, two clinical experts in gynecology (MNA, SG) independently examined the full EHR for each patient in the CUIMC sample, including clinical encounter notes, imaging reports, and surgical notes. We defined a gold-standard case as a patient with mention of a formal diagnosis of endometriosis in her chart and defined all other patients in the sample as controls. Any initial disagreement among reviewers was discussed until consensus was reached. To assess inter-rater reliability among the two reviewers, we calculated percent initial agreement and Cohen’s kappa (to account for the possibility of agreement among experts due to chance).^21,22^

To assess the performance of each phenotype definition, we computed precision (also called positive predictive value, PPV), recall (also known as sensitivity), and F1 score (the harmonic mean of precision and recall). We also computed concordance among patients identified by different cohort definitions in the sample set by calculating a Fleiss’ kappa score (to measure global agreement among all five phenotypes) and percent agreement and Cohen’s kappa scores (to measure pairwise agreement between phenotypes).

### Supplementary Results

Upon review, clinical experts classified 85% of charts in the stratified sample as gold-standard cases of endometriosis and 15% as controls. Concordance was high among chart reviewers (99% initial agreement; Cohen’s kappa = 0.96). We also found high levels of precision among all five cohort definitions (PPV range: 0.84-0.96; see **Supplementary Table S12**).

In contrast, complete agreement across all five cohort definitions was only found among 16% of cases in our stratified random sample (Fleiss’ kappa = 0.13). Pairwise percent agreement among cohort definitions varied widely from 38% to 81% (mean=58%, SD=16%; see **Supplementary Table S13**). Similarly, pairwise Cohen’s kappa scores ranged substantially from –0.18 to 0.62 (mean = 0.17, SD = 0.29; see **Supplementary Table S14**).

Close inspection of our chart review sample revealed nine (9%) cases where only the phenotype for Cohort A correctly identified patients with endometriosis and eight (8%) cases where only the phenotype for Cohort E correctly identified patients with endometriosis. There were six (6%) cases where only Cohorts C, D, and E (but not Cohorts A and B) correctly classified the patient as having endometriosis. We found no cases in which only Cohort B, only Cohort C, or only Cohort D correctly identified a patient with a diagnosis of endometriosis.

### Supplementary Discussion

Expert chart review confirmed that the majority of patients identified by at least one of the cohort definitions were true endometriosis cases. Moreover, while there was near perfect agreement among chart reviewers as to which patients were cases or controls, there was substantial lack of concordance among our five phenotype definitions. Notably, when assessed individually, each definition demonstrated very high precision (PPV range: 0.84-0.96).

Due to our desire to minimize false positives given the diagnostic nature of our study, we consider precision to be the primary performance metric for phenotype evaluation. Nonetheless, we recognize that chart review revealed recall levels that spanned high, moderate, and low values. Given that F1 scores represent the tradeoff between precision and recall, accordingly, most F1 values ranged from moderate to high performance.

Upon closer examination, we also found a lack of shared overlap in the women that comprised each cohort. This is demonstrated by poor concordance as measured by percent agreement and kappa scores. Nonetheless, two or more cohort definitions commonly identified the same women as having endometriosis, there was simply a lack of consensus across all five definitions. This is illustrated by the relatively small number of women in our sample that were identified solely by a single phenotype definition (eight in Cohort A, nine in Cohort E, zero in all others). Unsurprisingly, as all inclusion criteria for Cohort C are incorporated in the phenotype definition for Cohort D and all inclusion criteria for Cohort D are part of the phenotype definition for Cohort E, any patient identified by Cohort C was included in Cohort D and any patient identified by Cohort D was included in Cohort E.

**Table S1.** Participant Characteristics, Merative Commercial Claims and Encounters (CCAE) Dataset.

| **Characteristic** | **Cohort A**<br>(N = 142,833) | **Cohort B**<br>(N = 76,081) | **Cohort C**<br>(N = 173,105) | **Cohort D**<br>(N = 177,215) | **Cohort E**<br>(N = 182,908) |
| --- | --- | --- | --- | --- | --- |
| **Age Group** |  |  |  |  |  |
| 15-19 years | 2,180 (1.5%) | 2,263 (3.0%) | 4,493 (2.6%) | 4,560 (2.6%) | 4,657 (2.5%) |
| 20-24 years | 7,516 (5.3%) | 6,943 (9.1%) | 13,688 (7.9%) | 14,030 (7.9%) | 14,369 (7.9%) |
| 25-29 years | 10,995 (7.7%) | 8,000 (11%) | 16,891 (9.8%) | 17,541 (9.9%) | 18,149 (9.9%) |
| 30-34 years | 19,607 (14%) | 13,142 (17%) | 27,236 (16%) | 27,950 (16%) | 28,789 (16%) |
| 35-39 years | 28,810 (20%) | 16,739 (22%) | 36,520 (21%) | 37,367 (21%) | 38,473 (21%) |
| 40-44 years | 37,230 (26%) | 16,631 (22%) | 40,630 (23%) | 41,437 (23%) | 42,880 (23%) |
| 45-49 years | 36,495 (26%) | 12,363 (16%) | 33,647 (19%) | 34,330 (19%) | 35,591 (19%) |
| **Race** |  |  |  |  |  |
| Unknown | 142,833 (100%) | 76,081 (100%) | 173,105 (100%) | 177,215 (100%) | 182,908 (100%) |
| **Ethnicity** |  |  |  |  |  |
| Unknown | 142,833 (100%) | 76,081 (100%) | 173,105 (100%) | 177,215 (100%) | 182,908 (100%) |

**Table S2.** Participant Characteristics, Merative Medicaid (MDCD) Dataset.

| **Characteristic** | **Cohort A**<br>(N = 34,395) | **Cohort B**<br>(N = 24,466) | **Cohort C**<br>(N = 59,473) | **Cohort D**<br>(N = 60,504) | **Cohort E**<br>(N = 61,427) |
| --- | --- | --- | --- | --- | --- |
| **Age Group** |  |  |  |  |  |
| 15-19 years | 1,431 (4.2%) | 1,530 (6.3%) | 3,124 (5.3%) | 3,165 (5.2%) | 3,208 (5.2%) |
| 20-24 years | 2,585 (7.5%) | 2,596 (11%) | 5,324 (9.0%) | 5,446 (9.0%) | 5,523 (9.0%) |
| 25-29 years | 5,801 (17%) | 5,036 (21%) | 10,498 (18%) | 10,715 (18%) | 10,890 (18%) |
| 30-34 years | 7,251 (21%) | 5,781 (24%) | 13,357 (22%) | 13,628 (23%) | 13,828 (23%) |
| 35-39 years | 7,330 (21%) | 4,841 (20%) | 12,437 (21%) | 12,635 (21%) | 12,827 (21%) |
| 40-44 years | 6,126 (18%) | 3,142 (13%) | 9,377 (16%) | 9,491 (16%) | 9,644 (16%) |
| 45-49 years | 3,871 (11%) | 1,540 (6.3%) | 5,356 (9.0%) | 5,424 (9.0%) | 5,507 (9.0%) |
| **Race** |  |  |  |  |  |
| Black / African American | 7,133 (21%) | 5,287 (22%) | 13,603 (23%) | 13,847 (23%) | 14,106 (23%) |
| White | 22,717 (66%) | 15,895 (65%) | 38,037 (64%) | 38,670 (64%) | 39,218 (64%) |
| Unknown | 4,545 (13%) | 3,284 (13%) | 7,833 (13%) | 7,987 (13%) | 8,103 (13%) |
| **Ethnicity** |  |  |  |  |  |
| Hispanic / Latina | 1,175 (3.4%) | 816 (3.3%) | 2,046 (3.4%) | 2,102 (3.5%) | 2,135 (3.5%) |
| Unknown | 33,220 (97%) | 23,650 (97%) | 57,427 (97%) | 58,402 (97%) | 59,292 (97%) |

**Table S3.** Participant Characteristics, Columbia University Irving Medical Center (CUIMC) EHR Dataset.

| **Characteristic** | **Cohort A**<br>(N = 1,046) | **Cohort B**<br>(N = 766) | **Cohort C**<br>(N = 1,592) | **Cohort D**<br>(N = 1,663) | **Cohort E**<br>(N = 1,738) |
| --- | --- | --- | --- | --- | --- |
| **Age Group** |  |  |  |  |  |
| 15-19 years | 32 (3.1%) | 28 (3.7%) | 44 (2.8%) | 44 (2.6%) | 44 (2.5%) |
| 20-24 years | 38 (3.6%) | 35 (4.6%) | 62 (3.9%) | 69 (4.1%) | 72 (4.1%) |
| 25-29 years | 55 (5.3%) | 68 (8.9%) | 137 (8.6%) | 145 (8.7%) | 151 (8.7%) |
| 30-34 years | 104 (9.9%) | 111 (14%) | 205 (13%) | 212 (13%) | 228 (13%) |
| 35-39 years | 204 (20%) | 177 (23%) | 359 (23%) | 373 (22%) | 383 (22%) |
| 40-44 years | 341 (33%) | 201 (26%) | 409 (26%) | 422 (25%) | 442 (25%) |
| 45-49 years | 272 (26%) | 146 (19%) | 376 (24%) | 398 (24%) | 418 (24%) |
| **Race** |  |  |  |  |  |
| Asian | 55 (5.3%) | 23 (3.0%) | 54 (3.4%) | 60 (3.6%) | 62 (3.6%) |
| Black / African American | 168 (16%) | 128 (17%) | 259 (16%) | 272 (16%) | 278 (16%) |
| Native American / Alaska Native | 1 (<0.1%) | 2 (0.3%) | 4 (0.3%) | 4 (0.2%) | 4 (0.2%) |
| Native Hawaiian / Pacific Islander | 6 (0.6%) | 1 (0.1%) | 4 (0.3%) | 4 (0.2%) | 5 (0.3%) |
| White | 357 (34%) | 239 (31%) | 506 (32%) | 535 (32%) | 564 (32%) |
| Unknown | 459 (44%) | 373 (49%) | 765 (48%) | 788 (47%) | 825 (47%) |
| **Ethnicity** |  |  |  |  |  |
| Hispanic / Latina | 371 (35%) | 335 (44%) | 651 (41%) | 674 (41%) | 689 (40%) |
| Not Hispanic / Latina | 478 (46%) | 307 (40%) | 653 (41%) | 686 (41%) | 722 (42%) |
| Unknown | 197 (19%) | 124 (16%) | 288 (18%) | 303 (18%) | 327 (19%) |

**Table S4.** Patient Set Overlap Among Study Cohorts, All Datasets.

| **Cohort Combinations** | **CCAE**<br>**(N = 236,594)** | **Optum EHR**<br>**(N = 181,743)** | **MDCD**<br>**(N = 70,426)** | **CUIMC EHR**<br>**(N = 2,285)** |
| --- | --- | --- | --- | --- |
| Cohort A only | 53,686 (23%) | 42,375 (23%) | 8,999 (13%) | 547 (24%) |
| Cohort B only | 3,635 (1.5%) | 4,038 (2.2%) | 676 (1.0%) | 56 (2.5%) |
| Cohorts A and E | 2,058 (0.9%) | 1,511 (0.8%) | 247 (0.4%) | 19 (0.8%) |
| Cohorts D and E | 2,370 (1.0%) | 3,240 (1.8%) | 733 (1.0%) | 47 (2.1%) |
| Cohorts A, D, and E | 1,740 (0.7%) | 1,700 (0.9%) | 298 (0.4%) | 24 (1.1%) |
| Cohorts C, D, and E | 60,936 (26%) | 52,466 (29%) | 24,466 (35%) | 687 (30%) |
| Cohorts A, C, D, and E | 36,088 (15%) | 28,172 (16%) | 10,541 (15%) | 139 (6.1%) |
| Cohorts B, C, D, and E | 26,820 (11%) | 21,302 (12%) | 10,156 (14%) | 449 (20%) |
| Cohorts A-E | 49,261 (21%) | 26,939 (15%) | 14,310 (20%) | 317 (14%) |

**Table S5.** Patient Race and Ethnicity Among Cohort Intersections, Optum EHR Dataset.

| Characteristic | Cohort<br>A only<br>(N=<br>42,375) | Cohort<br>B only<br>(N=<br>4,038) | Cohorts<br>A and E<br>(N=<br>1,511) | Cohort<br>D and E<br>(N=<br>3,240) | Cohorts A,<br>D, E (N=<br>1,700) | Cohorts<br>C-E<br>(N=<br>52,466) | Cohorts<br>A, C-E<br>(N=<br>28,172) | Cohorts<br>B-E<br>(N=<br>21,302) | Cohorts<br>A-E<br>(N=<br>26,939) |
| --- | --- | --- | --- | --- | --- | --- | --- | --- | --- |
|  | N (%) |  |  |  |  |  |  |  |  |
| **Race** |  |  |  |  |  |  |  |  |  |
| Asian | 1,148 (3) | 86 (2) | 38 (3) | 58 (2) | 33 (2) | 903 (2) | 386 (1) | 495 (2) | 371 (1) |
| Black / African<br>American | 5,150 (12) | 421 (10) | 137 (9) | 292 (9) | 141 (8) | 6,173 (12) | 2,976 (11) | 2,899 (14) | 2,827 (10) |
| White | 33,249 (78) | 3,130 (78) | 1,234 (82) | 2,585 (80) | 1,427 (84) | 41,550 (79) | 23,206 (82) | 16,293 (76) | 22,304 (83) |
| Unknown | 2,828 (7) | 401 (10) | 102 (7) | 305 (9) | 99 (6) | 3,840 (7) | 1,604 (6) | 1,615 (8) | 1,437 (5) |
| **Ethnicity** |  |  |  |  |  |  |  |  |  |
| Hispanic /<br>Latina | 2,880 (7) | 236 (6) | 68 (5) | 200 (6) | 90 (5) | 3,388 (7) | 1,729 (6) | 1,710 (8) | 1,693 (6) |
| Not Hispanic /<br>Latina | 36,166 (85) | 3,355 (83) | 1,336 (88) | 2,670 (82) | 1,477 (87) | 45,335 (86) | 24,881 (88) | 17,989 (84) | 23,724 (88) |
| Unknown | 3,329 (8) | 447 (11) | 107 (7) | 370 (11) | 133 (8) | 3,743 (7) | 1,562 (6) | 1,603 (8) | 1,522 (6) |

**Table S6.** Patient Race and Ethnicity Among Cohort Intersections, MDCD Dataset.

| **Characteristic** | **Cohort**<br>**A only**<br>**(N=**<br>**8,999)** | **Cohort**<br>**B only**<br>**(N=**<br>**676)** | **Cohorts**<br>**A and E**<br>**(N=**<br>**247)** | **Cohort**<br>**D and E**<br>**(N=**<br>**733)** | **Cohorts**<br>**A, D, E**<br>**(N=**<br>**298)** | **Cohorts**<br>**C-E**<br>**(N=**<br>**24,466)** | **Cohorts**<br>**A, C-E**<br>**(N=**<br>**10,541)** | **Cohorts B-**<br>**E**<br>**(N=**<br>**10,156)** | **Cohorts**<br>**A-E**<br>**(N=**<br>**14,310)** |
| --- | --- | --- | --- | --- | --- | --- | --- | --- | --- |
|  | **N (%)** |  |  |  |  |  |  |  |  |
| **Race** |  |  |  |  |  |  |  |  |  |
| Black / African American | 2,137 (24) | 187 (28) | 72 (29) | 186 (25) | 58 (19) | 6,140 (25) | 2,176 (21) | 2,597 (26) | 2,690 (19) |
| White | 5,645 (63) | 402 (59) | 146 (59) | 440 (60) | 193 (65) | 15,176 (62) | 6,966 (66) | 6,128 (60) | 9,767 (68) |
| Unknown | 1,217 (14) | 87 (13) | 29 (12) | 107 (15) | 47 (16) | 3,150 (13) | 1,399 (13) | 1,431 (14) | 1,853 (13) |
| **Ethnicity** |  |  |  |  |  |  |  |  |  |
| Hispanic / Latina | 358 (4) | 22 (3) | 11 (5) | 39 (5) | 17 (6) | 870 (4) | 360 (3) | 387 (4) | 429 (3) |
| Unknown | 8,641 (96) | 654 (97) | 236 (96) | 694 (95) | 281 (94) | 23,596 (96) | 10,181 (97) | 9,769 (96) | 13,881 (97) |

**Table S7.** Patient Race and Ethnicity Among Cohort Intersections, CUIMC EHR Dataset.

| Characteristic | Cohort<br>A only<br>(N=547) | Cohort<br>B only<br>(N=56) | Cohorts<br>A and E<br>(N=19) | Cohort<br>D and E<br>(N=47) | Cohorts<br>A, D, E<br>(N=24) | Cohorts<br>C-E<br>(N=687) | Cohorts<br>A, C-E<br>(N=139) | Cohorts<br>B-E<br>(N=449) | Cohorts<br>A-E<br>(N=317) |
| --- | --- | --- | --- | --- | --- | --- | --- | --- | --- |
|  | N (%) |  |  |  |  |  |  |  |  |
| **Race** |  |  |  |  |  |  |  |  |  |
| Asian | 36 (7) | ** | ** | ** | ** | 28 (4) | ** | 11 (2) | 12 (4) |
| Black / African<br>American | 90 (16) | ** | ** | 7 (15) | 6 (25) | 104 (15) | 27 (19) | 84 (19) | 44 (14) |
| Native American/<br>Alaska Native | ** | ** | ** | ** | ** | ** | ** | ** | ** |
| Native Hawaiian/<br>Pacific Islander | ** | ** | ** | ** | ** | ** | ** | ** | ** |
| White | 188 (34) | 20 (36) | 9 (47) | 20 (43) | 9 (38) | 218 (32) | 49 (35) | 137 (31) | 102 (32) |
| Unknown | 231 (42) | 30 (54) | 7 (37) | 17 (36) | ** | 334 (49) | 58 (42) | 216 (48) | 157 (50) |
| **Ethnicity** |  |  |  |  |  |  |  |  |  |
| Hispanic / Latina | 169 (31) | 12 (21) | ** | 19 (40) | ** | 261 (38) | 55 (40) | 195 (43) | 140 (44) |
| Not Hispanic /<br>Latina | 263 (48) | 25 (45) | 11 (58) | 19 (40) | 14 (58) | 287 (42) | 59 (42) | 176 (39) | 131 (41) |
| Unknown | 115 (21) | 19 (34) | ** | 9 (19) | ** | 139 (20) | 25 (18) | 78 (17) | 46 (15) |

\*\*Cell counts are suppressed to protect participant privacy due to small sample size.

**Table S8.** Healthcare Utilization, Optum EHR Dataset.

| **Encounter Type** | **Characteristic** | **Cohort A**<br>(N = 100,697) | **Cohort B**<br>(N=48,241) | **Cohort C**<br>(N = 128,879) | **Cohort D**<br>(N = 133,819) | **Cohort E**<br>(N = 139,368) |
| --- | --- | --- | --- | --- | --- | --- |
| Outpatient Visits | n (%) | 100,362 (100) | 47,755 (99) | 126,060 (98) | 130,889 (98) | 136,191 (98) |
|  | mean (SD) | 17.4 (16.7) | 20.5 (18.8) | 17.1 (17.6) | 16.8 (17.4) | 16.4 (17.3) |
|  | median (IQR) | 13 (6-23) | 15 (8-27) | 12 (5-23) | 12 (5-22) | 11 (5-22) |
| ER Visits | n (%) | 30,560 (30) | 19,738 (41) | 43,170 (34) | 44,118 (33) | 44,949 (32) |
|  | mean (SD) | 0.8 (3.2) | 1.4 (4.3) | 1 (3.9) | 1 (3.8) | 0.9 (3.8) |
|  | median (IQR) | 0 (0-1) | 0 (0-1) | 0 (0-1) | 0 (0-1) | 0 (0-1) |
| Inpatient Visits | n (%) | 45,459 (45) | 12,287 (26) | 32,564 (25) | 33,599 (25) | 34,830 (25) |
|  | mean (SD) | 0.6 (1.1) | 0.4 (1.8) | 0.4 (1.5) | 0.4 (1.5) | 0.4 (1.4) |
|  | median (IQR) | 0 (0-1) | 0 (0-1) | 0 (0-1) | 0 (0-1) | 0 (0-1) |

**Table S9.** Healthcare Utilization, CCAE Dataset.

| **Encounter Type** | **Characteristic** | **Cohort A**<br>(N = 142,833) | **Cohort B**<br>(N = 76,081) | **Cohort C**<br>(N = 173,105) | **Cohort D**<br>(N = 177,215) | **Cohort E**<br>(N = 182,908) |
| --- | --- | --- | --- | --- | --- | --- |
| Outpatient Visits | n (%) | 142,749 (100) | 76,057 (100) | 173,043 (100) | 177,146 (100) | 182,832 (100) |
|  | mean (SD) | 16.8 (13.5) | 18.3 (15.2) | 17.1 (14.4) | 17 (14.3) | 16.8 (14.3) |
|  | median (IQR) | 13 (8-21) | 14 (8-23) | 13 (8-21) | 13 (8-21) | 13 (8-21) |
| ER Visits | n (%) | 45,285 (32) | 27,532 (36) | 58,172 (34) | 59,206 (33) | 60,359 (33) |
|  | mean (SD) | 0.6 (2.5) | 0.8 (3.3) | 0.7 (2.8) | 0.7 (2.8) | 0.7 (2.8) |
|  | median (IQR) | 0 (0-1) | 0 (0-1) | 0 (0-1) | 0 (0-1) | 0 (0-1) |
| Inpatient Visits | n (%) | 62,273 (44) | 17,542 (23) | 44,470 (26) | 45,349 (26) | 46,864 (26) |
|  | mean (SD) | 0.5 (1) | 0.3 (1.4) | 0.3 (1.2) | 0.3 (1.2) | 0.3 (1.2) |
|  | median (IQR) | 0 (0-1) | 0 (0-0) | 0 (0-1) | 0 (0-1) | 0 (0-1) |

**Table S10.** Healthcare Utilization, MDCD Dataset.

| **Encounter Type** | **Characteristic** | **Cohort A**<br>(N = 34,395) | **Cohort B**<br>(N = 24,466) | **Cohort C**<br>(N = 59,473) | **Cohort D**<br>(N = 60,504) | **Cohort E**<br>(N = 61,427) |
| --- | --- | --- | --- | --- | --- | --- |
| Outpatient Visits | n (%) | 34,335 (100) | 24,420 (100) | 59,300 (100) | 60,325 (100) | 61,233 (100) |
|  | mean (SD) | 22.5 (23) | 23.3 (22.4) | 22.3 (23) | 22.1 (22.9) | 22 (22.9) |
|  | median (IQR) | 17 (11-27) | 18 (11-29) | 17 (10-27) | 17 (10-27) | 17 (10-27) |
| ER Visits | n (%) | 24,150 (70) | 18,812 (77) | 43,595 (73) | 44,125 (73) | 44,551 (73) |
|  | mean (SD) | 2.8 (5.1) | 3.9 (6.9) | 3.3 (5.9) | 3.3 (5.9) | 3.2 (5.9) |
|  | median (IQR) | 1 (0-4) | 2 (1-5) | 2 (0-4) | 2 (0-4) | 2 (0-4) |
| Inpatient Visits | n (%) | 17,548 (51) | 8,334 (34) | 21,205 (36) | 21,446 (35) | 21,739 (35) |
|  | mean (SD) | 0.8 (1.2) | 0.6 (1.4) | 0.6 (1.3) | 0.6 (1.3) | 0.5 (1.3) |
|  | median (IQR) | 1 (0-1) | 0 (0-1) | 0 (0-1) | 0 (0-1) | 0 (0-1) |

**Table S11.** Healthcare Utilization, CUIMC EHR Dataset.

| **Encounter Type** | **Characteristic** | **Cohort A**<br>(N = 1,046) | **Cohort B**<br>(N = 766) | **Cohort C**<br>(N = 1,592) | **Cohort D**<br>(N = 1,663) | **Cohort E**<br>(N = 1,738) |
| --- | --- | --- | --- | --- | --- | --- |
| Outpatient Visits | n (%) | 833 | 494 | 975 | 1013 | 1060 |
|  | mean (SD) | 7.3 (12.2) | 5.4 (10.7) | 5.1 (10.6) | 5 (10.5) | 4.9 (10.3) |
|  | median (IQR) | 5 (1-10) | 2 (0-7) | 2 (0-7) | 2 (0-6) | 2 (0-6) |
| ER Visits | n (%) | 218 | 235 | 442 | 454 | 462 |
|  | mean (SD) | 0.4 (2) | 0.7 (2.3) | 0.6 (2.2) | 0.6 (2.2) | 0.6 (2.2) |
|  | median (IQR) | 0 (0-0) | 0 (0-1) | 0 (0-1) | 0 (0-1) | 0 (0-1) |
| Inpatient Visits | n (%) | 196 | 48 | 145 | 148 | 155 |
|  | mean (SD) | 0.2 (0.2) | 0.1 (0.5) | 0.1 (0.3) | 0.1 (0.3) | 0.1 (0.3) |
|  | median (IQR) | 0 (0-0) | 0 (0-0) | 0 (0-0) | 0 (0-0) | 0 (0-0) |

**Table S12.** Classification Report for Endometriosis Cohort Definitions.

|  | Cohort A | Cohort B | Cohort C | Cohort D | Cohort E |
| --- | --- | --- | --- | --- | --- |
| **precision (PPV)** | 0.96 | 0.93 | 0.84 | 0.84 | 0.84 |
| **recall (sensitivity)** | 0.64 | 0.32 | 0.48 | 0.67 | 0.89 |
| **F1 score** | 0.77 | 0.47 | 0.61 | 0.75 | 0.86 |

*Abbreviations:* PPV, positive predictive value.

**Table S13.** Pairwise Percent Agreement Between Cohorts.

|  |  | Cohort |  |  |  |  |
| --- | --- | --- | --- | --- | --- | --- |
|  |  | A | B | C | D | E |
| Cohort | A | 100% | 47% | 45% | 46% | 47% |
|  | B | 47% | 100% | 80% | 61% | 38% |
|  | C | 45% | 80% | 100% | 81% | 58% |
|  | D | 46% | 61% | 81% | 100% | 77% |
|  | E | 47% | 38% | 58% | 77% | 100% |

**Table S14.**
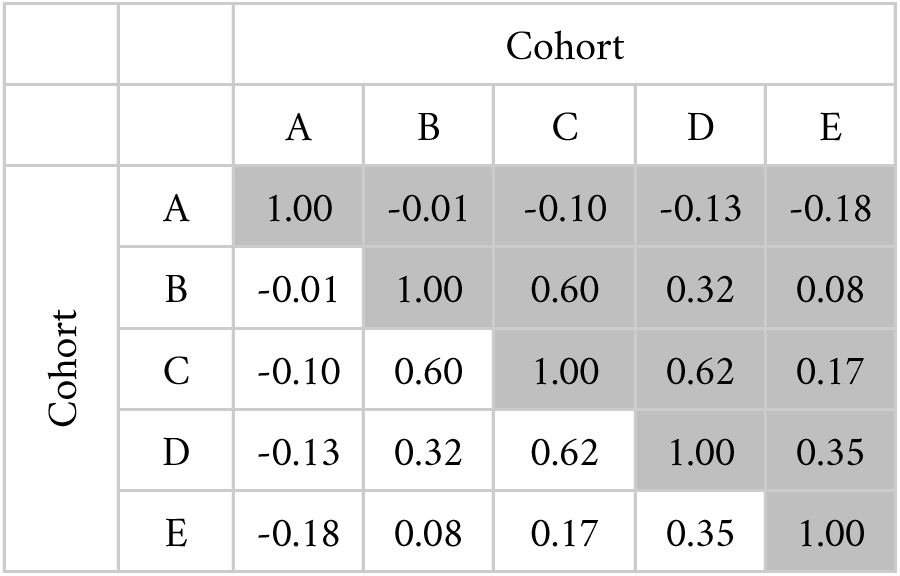
Pairwise Cohen’s Kappa Scores Between Cohorts.

|  |  | Cohort |  |  |  |  |
| --- | --- | --- | --- | --- | --- | --- |
|  |  | A | B | C | D | E |
| Cohort | A | 1.00 | -0.01 | -0.10 | -0.13 | -0.18 |
|  | B | -0.01 | 1.00 | 0.60 | 0.32 | 0.08 |
|  | C | -0.10 | 0.60 | 1.00 | 0.62 | 0.17 |
|  | D | -0.13 | 0.32 | 0.62 | 1.00 | 0.35 |
|  | E | -0.18 | 0.08 | 0.17 | 0.35 | 1.00 |

**Figure S1.**
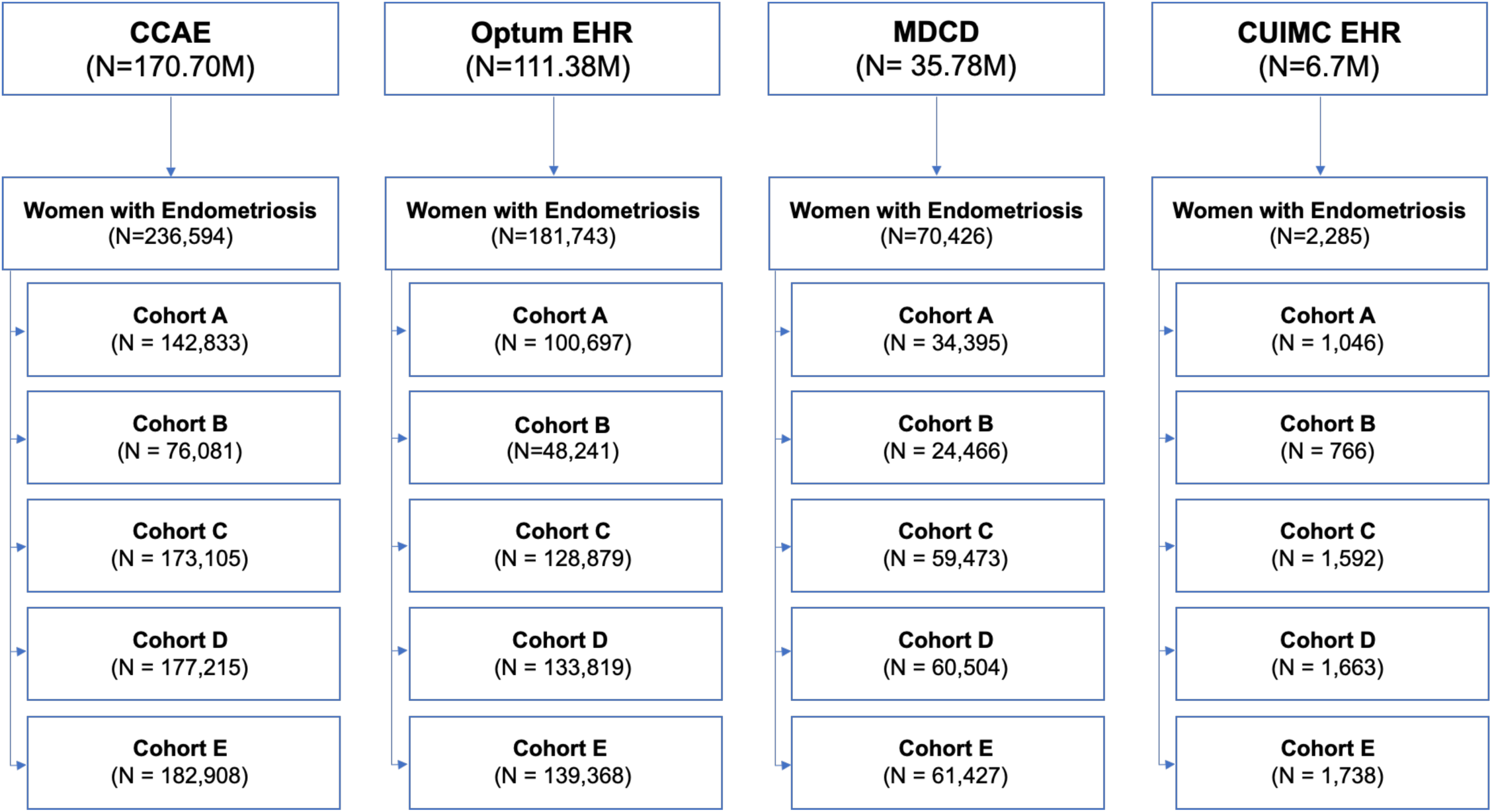
STROBE Flow Diagram of Women Diagnosed with Endometriosis Based on Surgery, Imaging, and/or Symptom Presentation in the CCAE, Optum EHR, MDCD, and CUIMC EHR Databases. Cohort A: Surgical confirmation phenotype; Cohort B: Imaging and guideline-recognized symptoms phenotype; Cohort C: Guideline-recognized symptoms only phenotype; Cohort D: Guideline-recognized symptoms and/or pelvic pain phenotype; Cohort E: Guideline-recognized symptoms, pelvic pain, and/or abdominal pain phenotype. *Abbreviations:* CCAE, Commercial Claims and Encounters. CUIMC, Columbia University Irving Medical Center. EHR, electronic health record. MDCD, Medicaid. STROBE: STrengthening the Reporting of OBservational studies in Epidemiology.

**Figure S2a-d.**
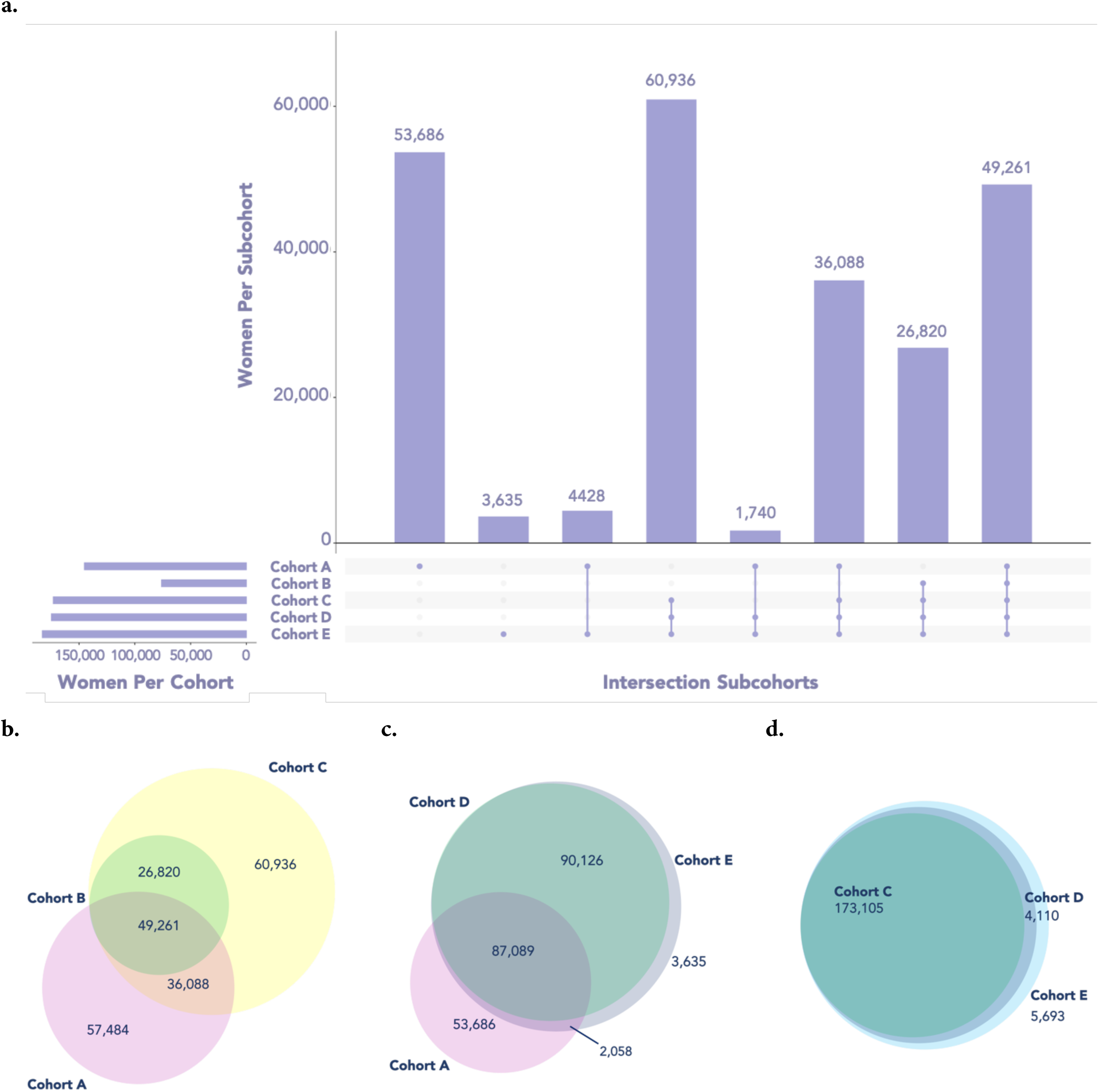
Patient Set Overlap Among Study Cohorts in the CCAE Dataset. a) UpSet plot illustrating intersections across all cohorts. b) Venn diagram of Cohorts A, B, and C. c) Venn diagram of Cohorts A, D, and E. d) Venn diagram of Cohorts C, D, and E. Cohort A: Surgical confirmation phenotype; Cohort B: Imaging and guideline-recognized symptoms phenotype; Cohort C: Guideline-recognized symptoms only phenotype; Cohort D: Guideline-recognized symptoms and/or pelvic pain phenotype; Cohort E: Guideline-recognized symptoms, pelvic pain, and/or abdominal pain phenotype. *Abbreviation:* CCAE, Commercial Claims and Encounters. EHR: electronic health record.

**Figure S3a-d.**
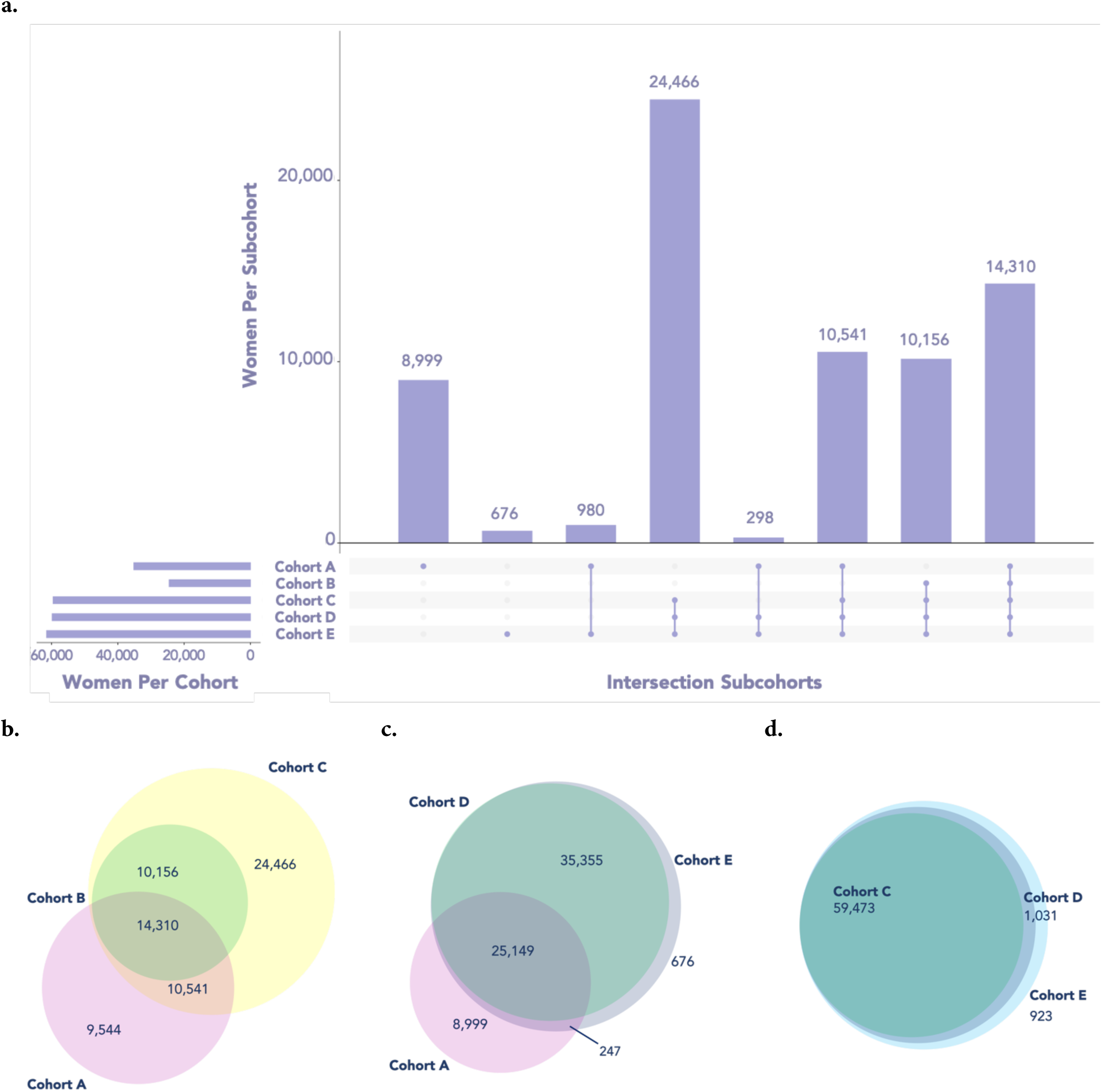
Patient Set Overlap Among Study Cohorts in MDCD Dataset. a) UpSet plot illustrating intersections across all cohorts. b) Venn diagram of Cohorts A, B, and C. c) Venn diagram of Cohorts A, D, and E. d) Venn diagram of Cohorts C, D, and E. Cohort A: Surgical confirmation phenotype; Cohort B: Imaging and guideline-recognized symptoms phenotype; Cohort C: Guideline-recognized symptoms only phenotype; Cohort D: Guideline-recognized symptoms and/or pelvic pain phenotype; Cohort E: Guideline-recognized symptoms, pelvic pain, and/or abdominal pain phenotype. *Abbreviations:* EHR, electronic health record. MDCD, Medicaid.

**Figure S4a-d.**
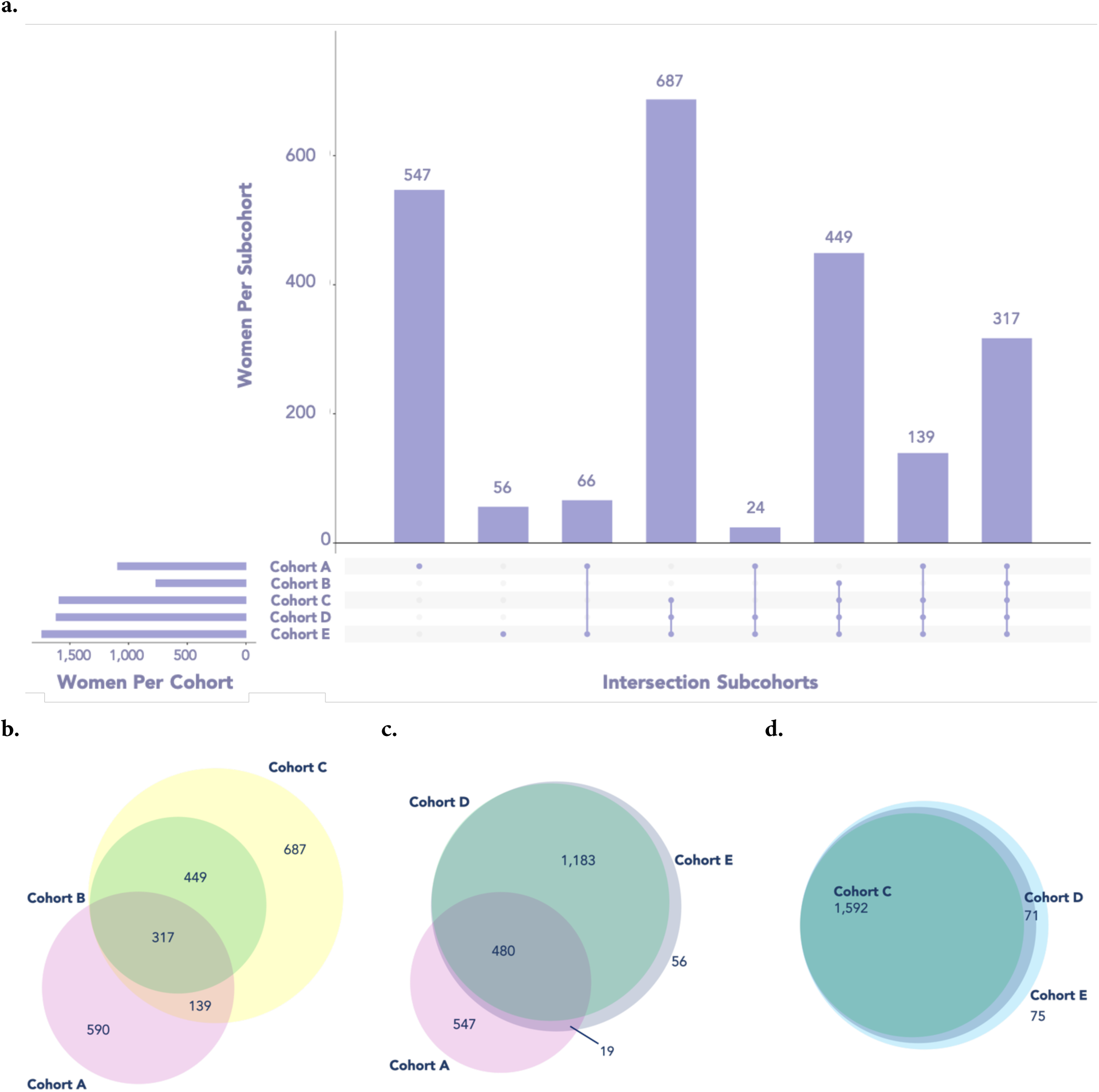
Patient Set Overlap Among Study Cohorts in CUIMC EHR Dataset. a) UpSet plot illustrating intersections across all cohorts. b) Venn diagram of Cohorts A, B, and C. c) Venn diagram of Cohorts A, D, and E. d) Venn diagram of Cohorts C, D, and E. Cohort A: Surgical confirmation phenotype; Cohort B: Imaging and guideline-recognized symptoms phenotype; Cohort C: Guideline-recognized symptoms only phenotype; Cohort D: Guideline-recognized symptoms and/or pelvic pain phenotype; Cohort E: Guideline-recognized symptoms, pelvic pain, and/or abdominal pain phenotype. *Abbreviations:* CUIMC, Columbia University Irving Medical Center. EHR, electronic health record.

**Figure S5a-d.**
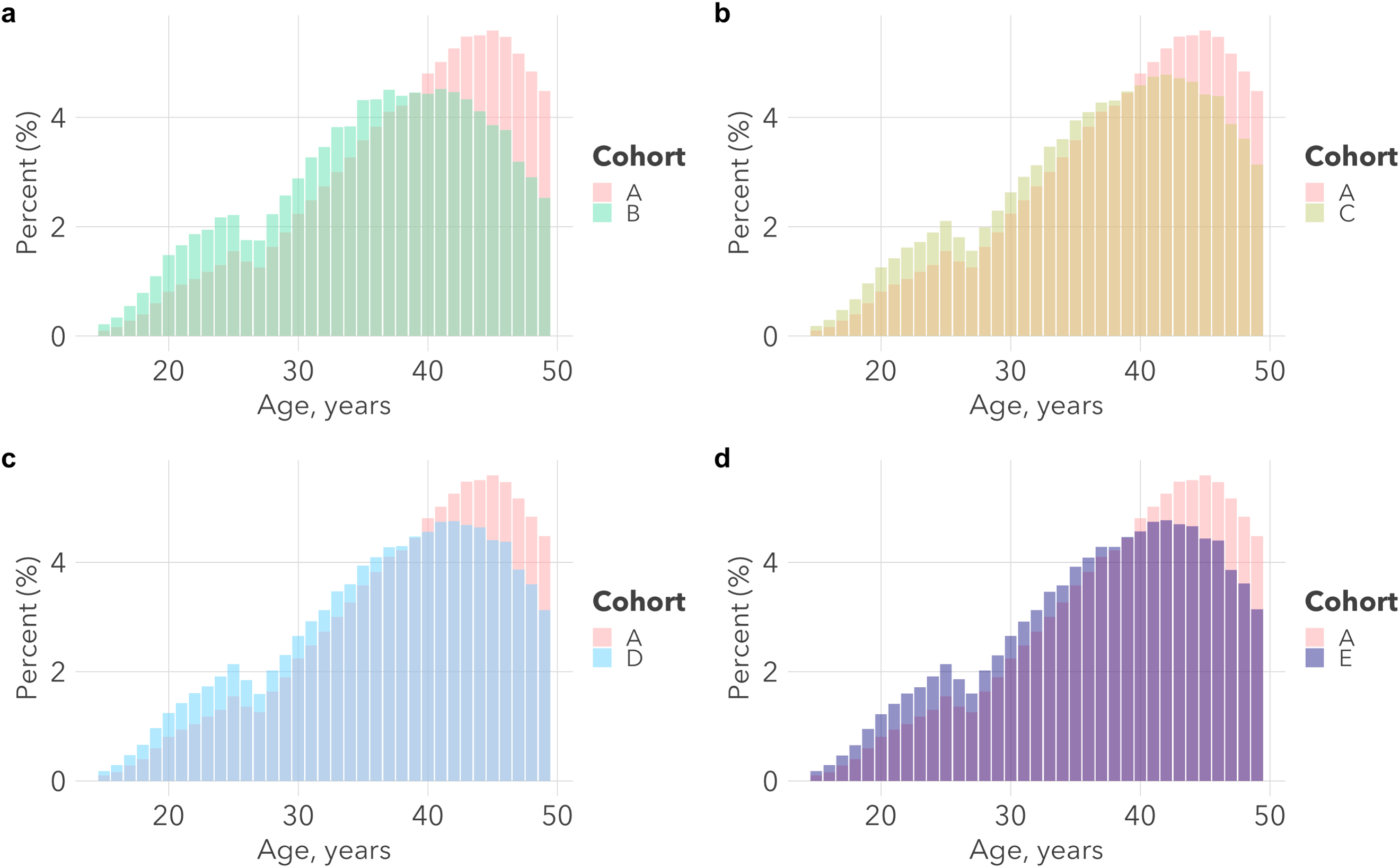
Age at Diagnosis of Endometriosis in the CCAE Dataset. On average, women in Cohort A were diagnosed with endometriosis at an older age than women in Cohorts B-E. a) Age at diagnosis among patients in Cohorts A and B. b) Age at diagnosis among patients in Cohorts A and C. c) Age at diagnosis among patients in Cohorts A and D. d) Age at diagnosis among patients in Cohorts A and E. Cohort A: Surgical confirmation phenotype; Cohort B: Imaging and guideline-recognized symptoms phenotype; Cohort C: Guideline-recognized symptoms only phenotype; Cohort D: Guideline-recognized symptoms and/or pelvic pain phenotype; Cohort E: Guideline-recognized symptoms, pelvic pain, and/or abdominal pain phenotype. *Abbreviations:* CCAE, Commercial Claims and Encounters.

**Figure S6a-d.**
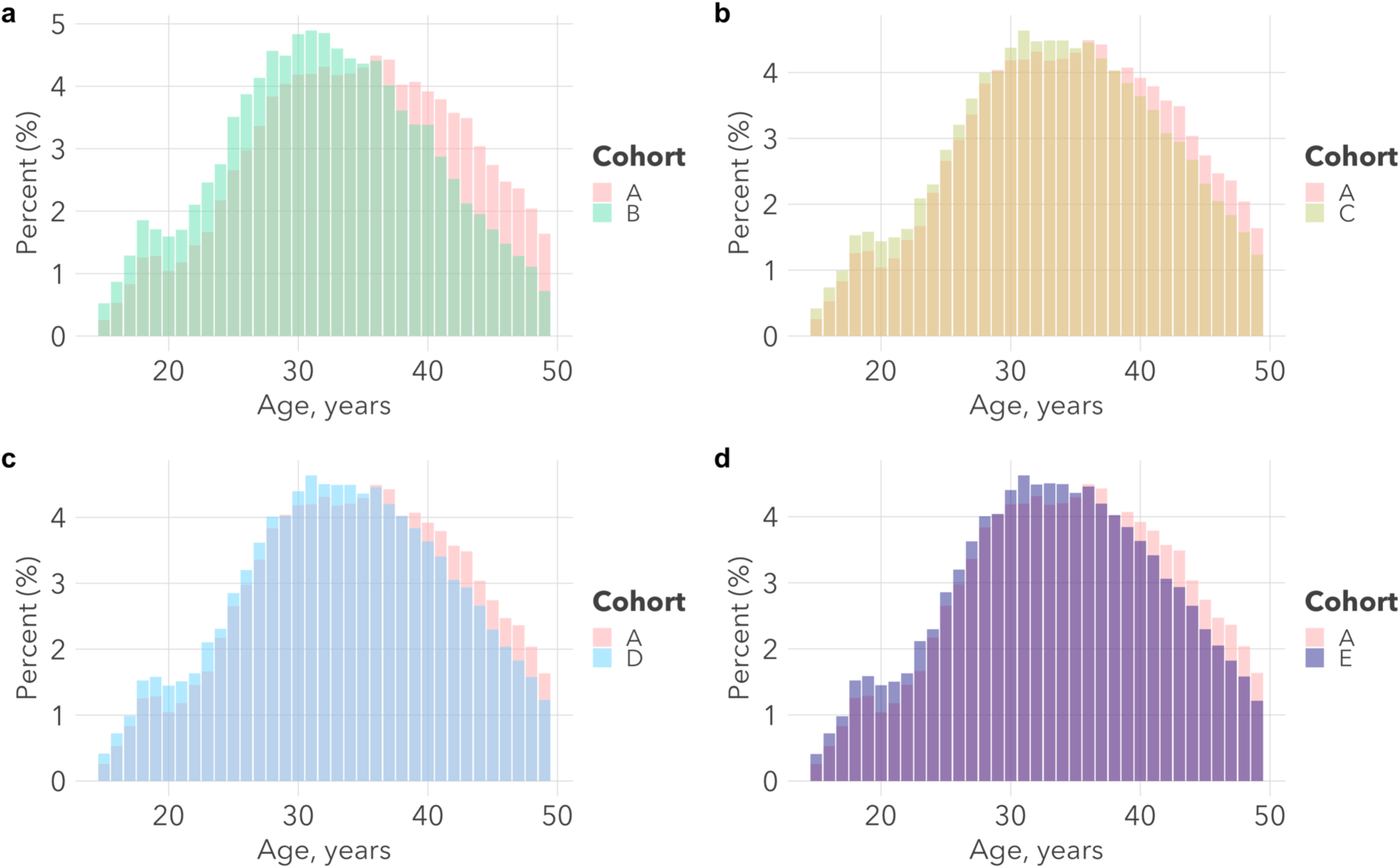
Age at Diagnosis of Endometriosis in the MDCD Dataset. On average, women in Cohort A were diagnosed with endometriosis at an older age than women in Cohorts B-E. a) Age at diagnosis among patients in Cohorts A and B. b) Age at diagnosis among patients in Cohorts A and C. c) Age at diagnosis among patients in Cohorts A and D. d) Age at diagnosis among patients in Cohorts A and E. Cohort A: Surgical confirmation phenotype; Cohort B: Imaging and guideline-recognized symptoms phenotype; Cohort C: Guideline-recognized symptoms only phenotype; Cohort D: Guideline-recognized symptoms and/or pelvic pain phenotype; Cohort E: Guideline-recognized symptoms, pelvic pain, and/or abdominal pain phenotype. *Abbreviations:* MDCD, Medicaid.

**Figure S7a-d.**
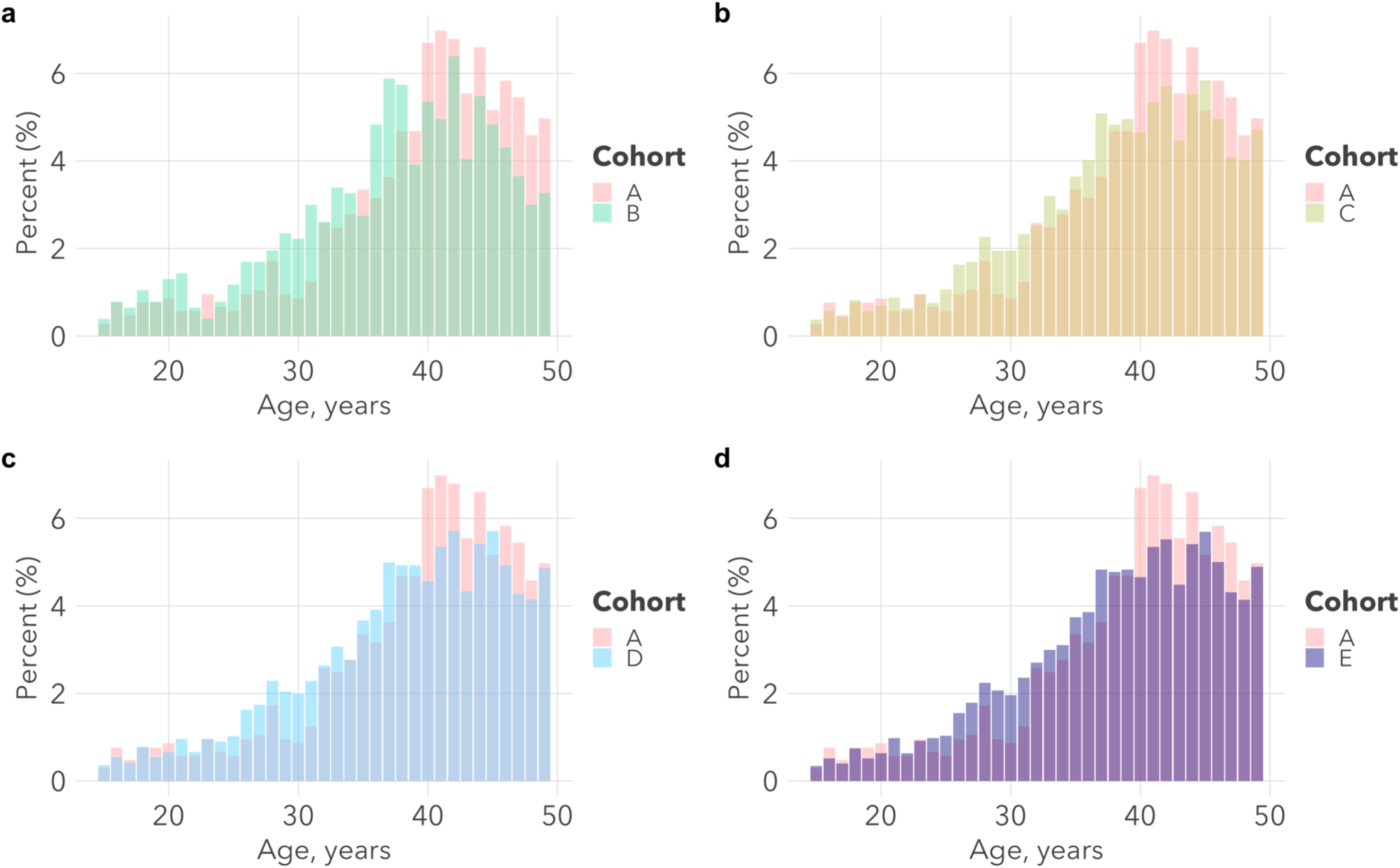
Age at Diagnosis of Endometriosis in the CUIMC EHR Dataset. On average, women in Cohort A were diagnosed with endometriosis at an older age than women in Cohorts B-E. a) Age at diagnosis among patients in Cohorts A and B. b) Age at diagnosis among patients in Cohorts A and C. c) Age at diagnosis among patients in Cohorts A and D. d) Age at diagnosis among patients in Cohorts A and E. Cohort A: Surgical confirmation phenotype; Cohort B: Imaging and guideline-recognized symptoms phenotype; Cohort C: Guideline-recognized symptoms only phenotype; Cohort D: Guideline-recognized symptoms and/or pelvic pain phenotype; Cohort E: Guideline-recognized symptoms, pelvic pain, and/or abdominal pain phenotype. *Abbreviations:* CUIMC, Columbia University Irving Medical Center. EHR, electronic health record.

**Figure S8a-d.**
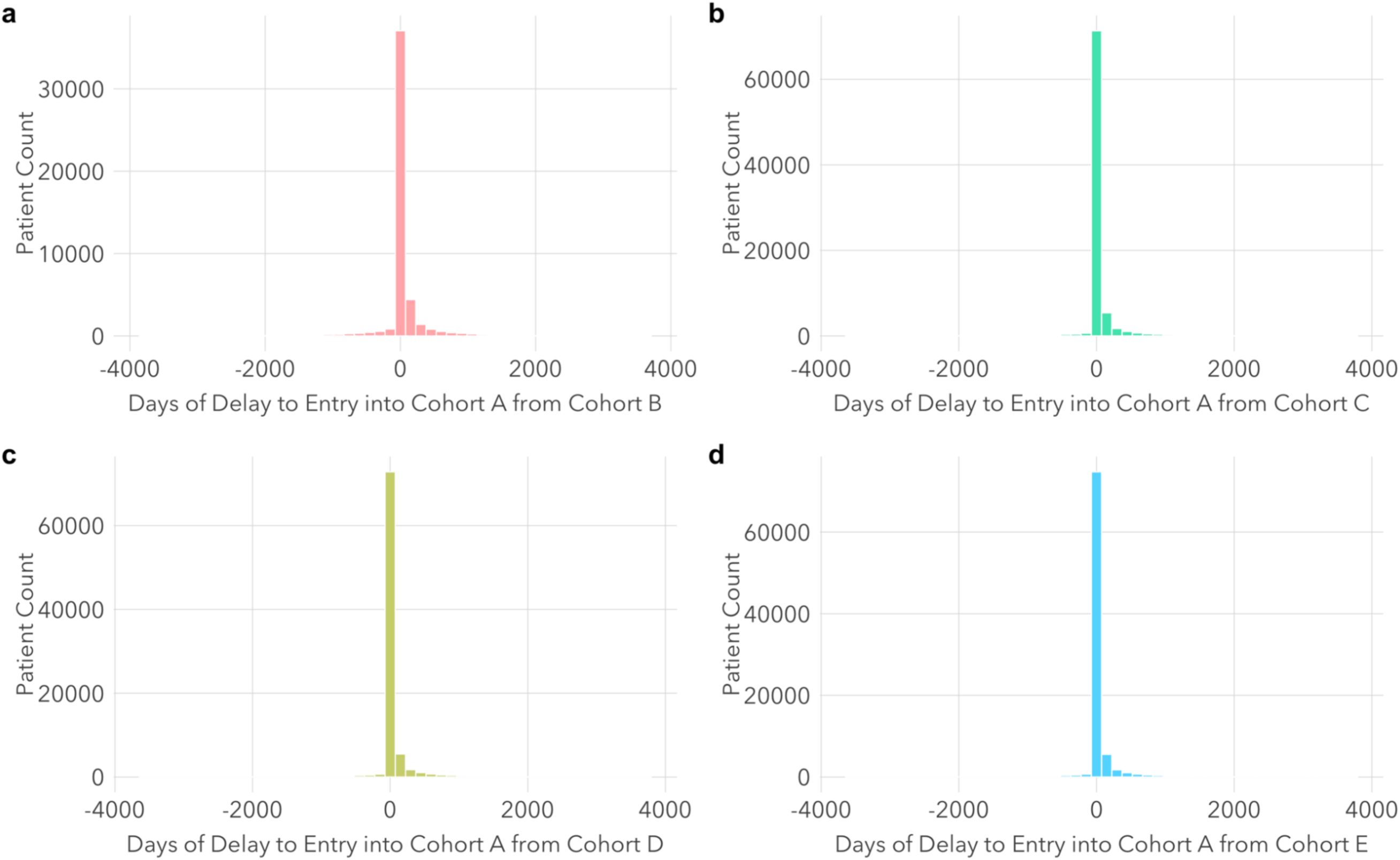
Differences in Cohort Entry Dates in the CCAE Dataset. Among most women identified by more than one phenotype definition, there was little to no difference in date of cohort entry (e.g., median difference in entry date between Cohort A and B = 0 days, interquartile range [IQR] = 0-40 days). a) Differences in cohort entry dates among patients identified by both cohort definitions A and B. b) Differences in cohort entry dates among patients identified by both cohort definitions A and C. c) Differences in cohort entry dates among patients identified by both cohort definitions A and D. d) Differences in cohort entry dates among patients identified by cohort definitions A and E. Cohort A: Surgical confirmation phenotype; Cohort B: Imaging and guideline-recognized symptoms phenotype; Cohort C: Guideline-recognized symptoms only phenotype; Cohort D: Guideline-recognized symptoms and/or pelvic pain phenotype; Cohort E: Guideline-recognized symptoms, pelvic pain, and/or abdominal pain phenotype. *Abbreviations:* CCAE, Commercial Claims and Encounters.

**Figure S9a-d.**
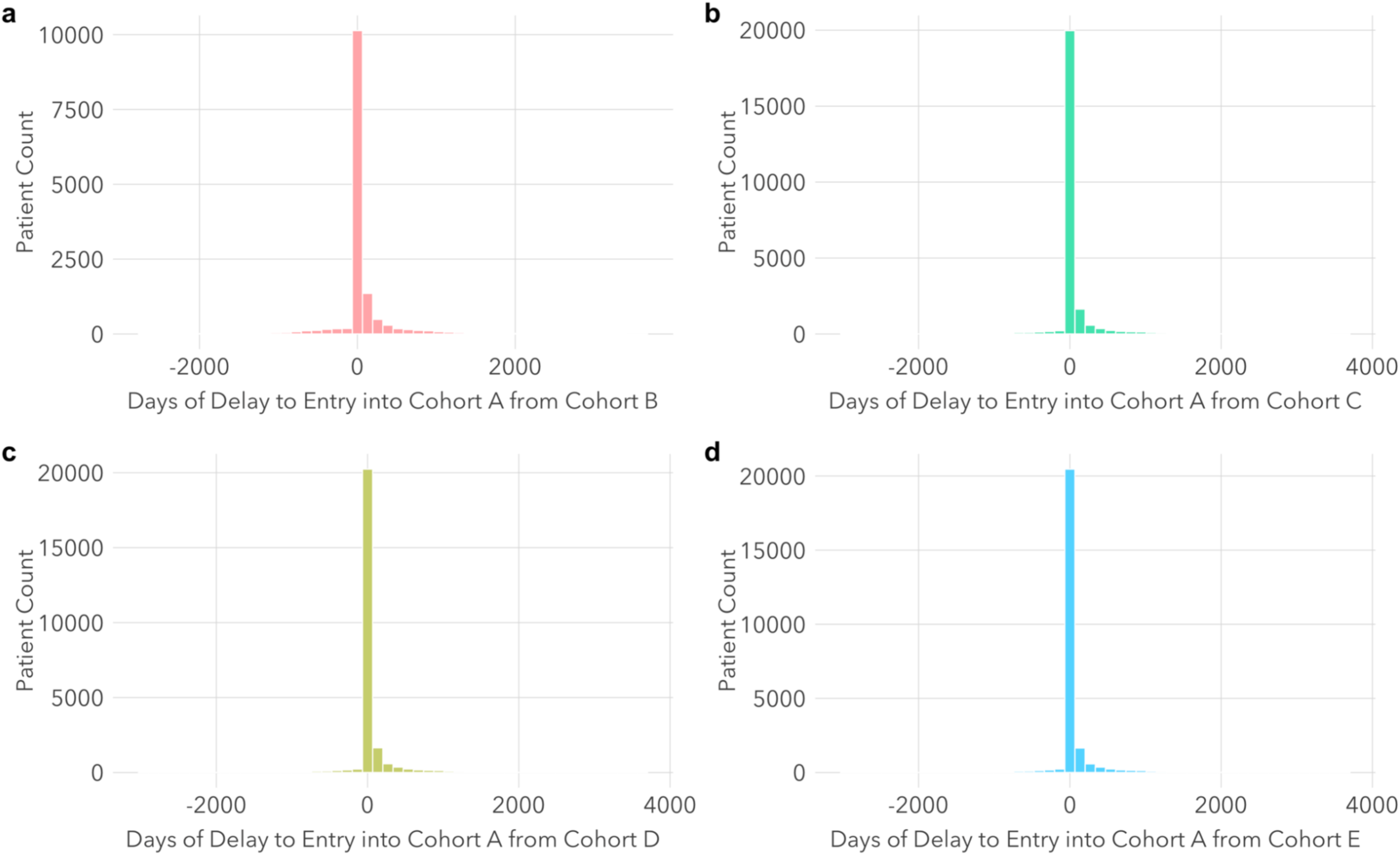
Differences in Cohort Entry Dates in the MDCD Dataset. Among most women identified by more than one phenotype definition, there was little to no difference in date of cohort entry (e.g., median difference in entry date between Cohort A and B = 0 days, interquartile range [IQR] = 0-47 days). a) Differences in cohort entry dates among patients identified by both cohort definitions A and B. b) Differences in cohort entry dates among patients identified by both cohort definitions A and C. c) Differences in cohort entry dates among patients identified by both cohort definitions A and D. d) Differences in cohort entry dates among patients identified by cohort definitions A and E. Cohort A: Surgical confirmation phenotype; Cohort B: Imaging and guideline-recognized symptoms phenotype; Cohort C: Guideline-recognized symptoms only phenotype; Cohort D: Guideline-recognized symptoms and/or pelvic pain phenotype; Cohort E: Guideline-recognized symptoms, pelvic pain, and/or abdominal pain phenotype. *Abbreviations:* MDCD, Medicaid.

**Figure S10a-d.**
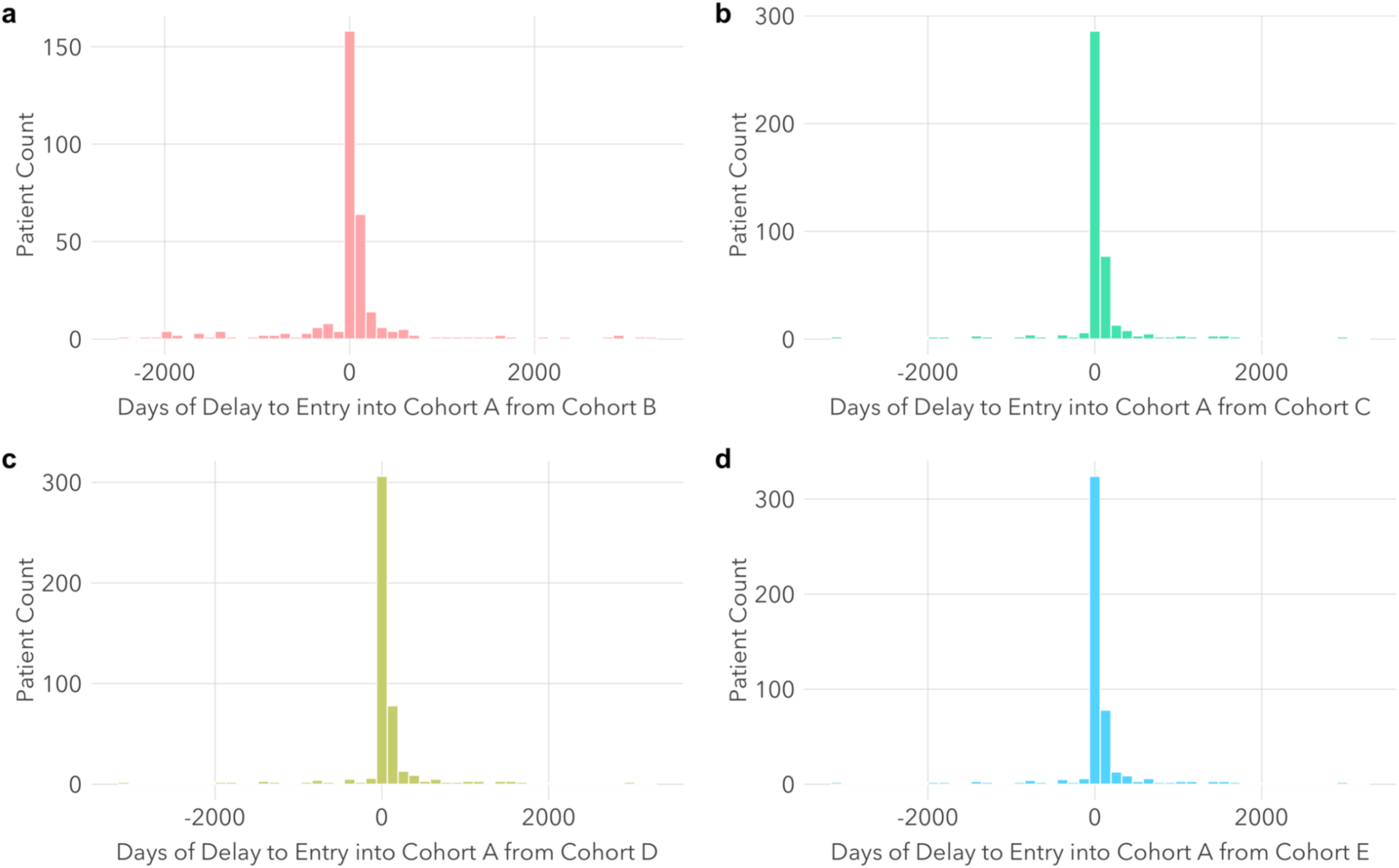
Differences in Cohort Entry Dates in the CUIMC EHR Dataset. Among most women identified by more than one phenotype definition, there was little to no difference in date of cohort entry (e.g., median difference in entry date between Cohort A and B = 19 days, interquartile range [IQR] = 0-91 days). a) Differences in cohort entry dates among patients identified by both cohort definitions A and B. b) Differences in cohort entry dates among patients identified by both cohort definitions A and C. c) Differences in cohort entry dates among patients identified by both cohort definitions A and D. d) Differences in cohort entry dates among patients identified by cohort definitions A and E. Cohort A: Surgical confirmation phenotype; Cohort B: Imaging and guideline-recognized symptoms phenotype; Cohort C: Guideline-recognized symptoms only phenotype; Cohort D: Guideline-recognized symptoms and/or pelvic pain phenotype; Cohort E: Guideline-recognized symptoms, pelvic pain, and/or abdominal pain phenotype. *Abbreviations:*CUIMC, Columbia University Irving Medical Center. EHR, electronic health record.

**Figure S11a-b.**
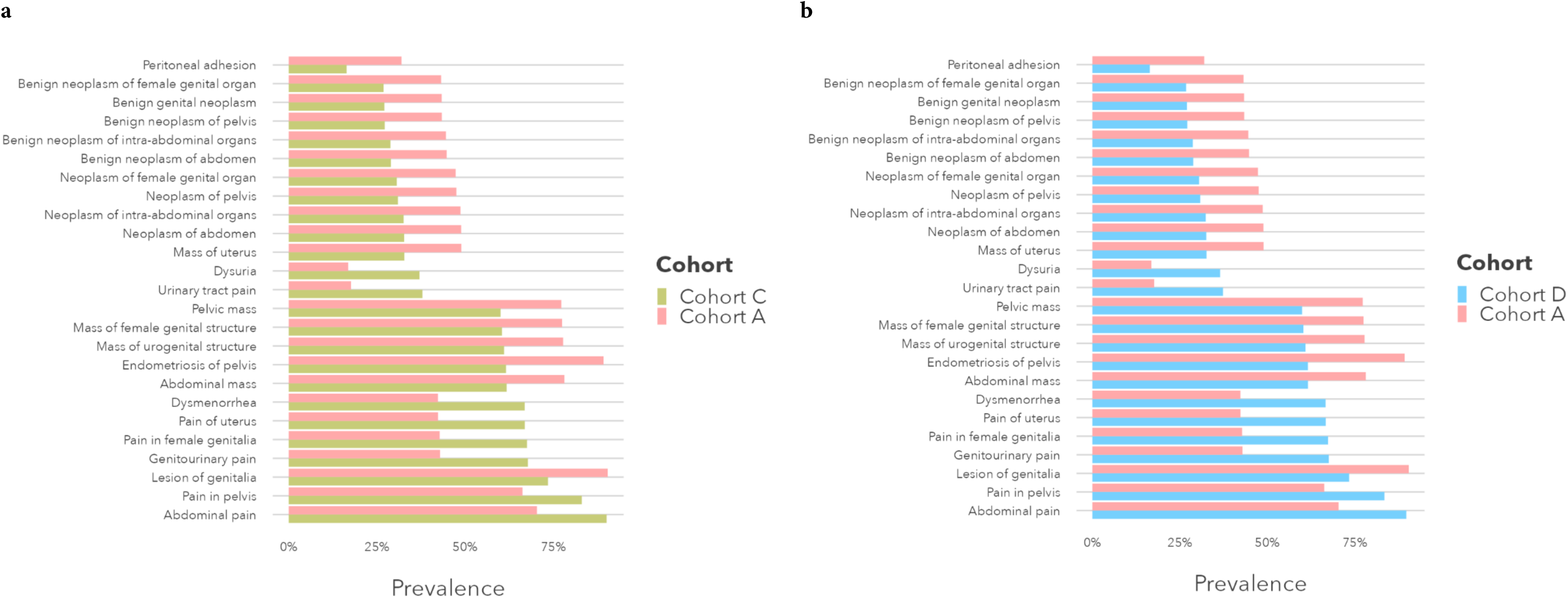
Condition Concepts in the Optum EHR Dataset with the Largest Differences in Prevalence between Endometriosis Cohorts Based on Surgical Diagnosis (Cohort A) and Symptoms-based Diagnosis (Cohorts C and D). Compared to Cohort A, localized pain (e.g., abdominal, pelvic, genitourinary), dysmenorrhea, and dysuria were consistently more common in Cohorts C and D. Lesions, masses, and neoplasms (e.g., abdominal, pelvic, uterine) were more common in Cohort A. Cohort A: Surgical confirmation phenotype; Cohort C: Guideline-recognized symptoms only phenotype; Cohort D: Guideline-recognized symptoms and/or pelvic pain phenotype. *Abbreviations:* EHR, electronic health record.

**Figure S12a-d.**
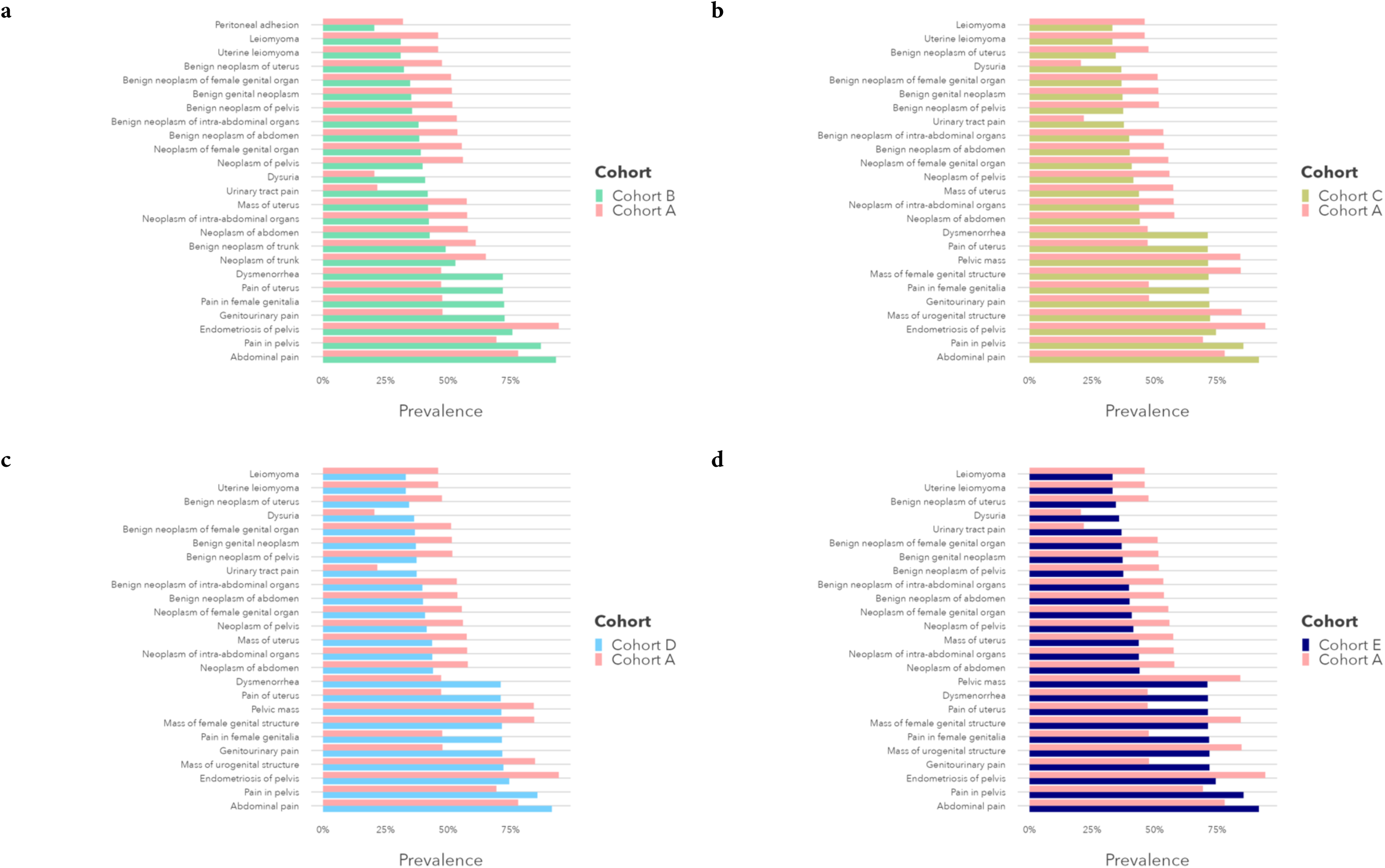
Condition Concepts in the CCAE Dataset with the Largest Differences in Prevalence Between Endometriosis Cohorts Based in Surgical Diagnosis (Cohort A) and Symptoms-based Diagnosis (Cohorts B-E). Cohort A: Surgical confirmation phenotype; Cohort B: Imaging and guideline-recognized symptoms phenotype; Cohort C: Guideline-recognized symptoms only phenotype; Cohort D: Guideline-recognized symptoms and/or pelvic pain phenotype; Cohort E: Guideline-recognized symptoms, pelvic pain, and/or abdominal pain phenotype. *Abbreviations:* CCAE, Commercial Claims and Encounters.

**Figure S13a-d.**
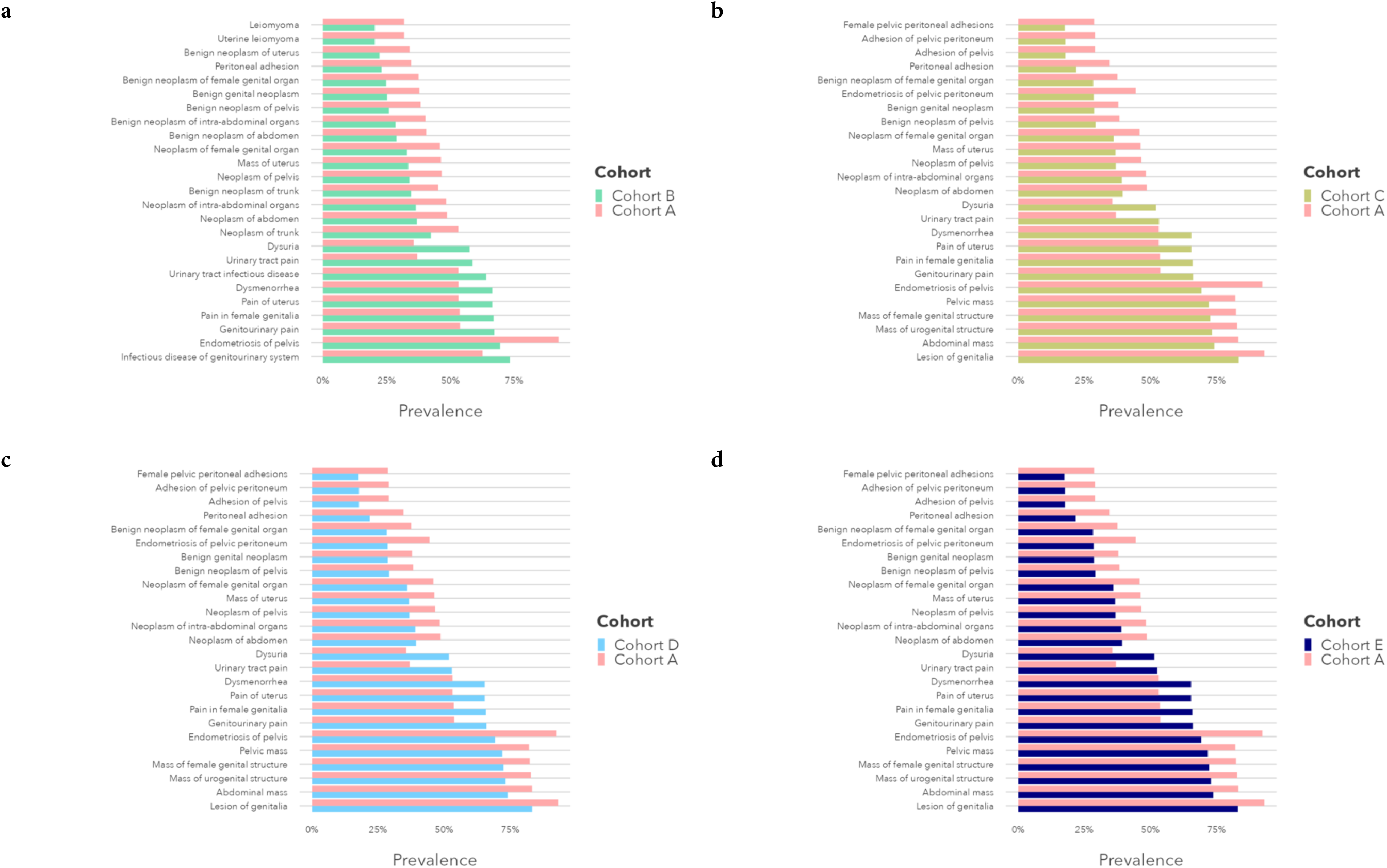
Condition Concepts in the MDCD Dataset with the Largest Differences in Prevalence Between Endometriosis Cohorts Based in Surgical Diagnosis (Cohort A) and Symptoms-based Diagnosis (Cohorts B-E). Cohort A: Surgical confirmation phenotype; Cohort B: Imaging and guideline-recognized symptoms phenotype; Cohort C: Guideline-recognized symptoms only phenotype; Cohort D: Guideline-recognized symptoms and/or pelvic pain phenotype; Cohort E: Guideline-recognized symptoms, pelvic pain, and/or abdominal pain phenotype. *Abbreviations:* MDCD, Medicaid.

**Figure S14a-d.**
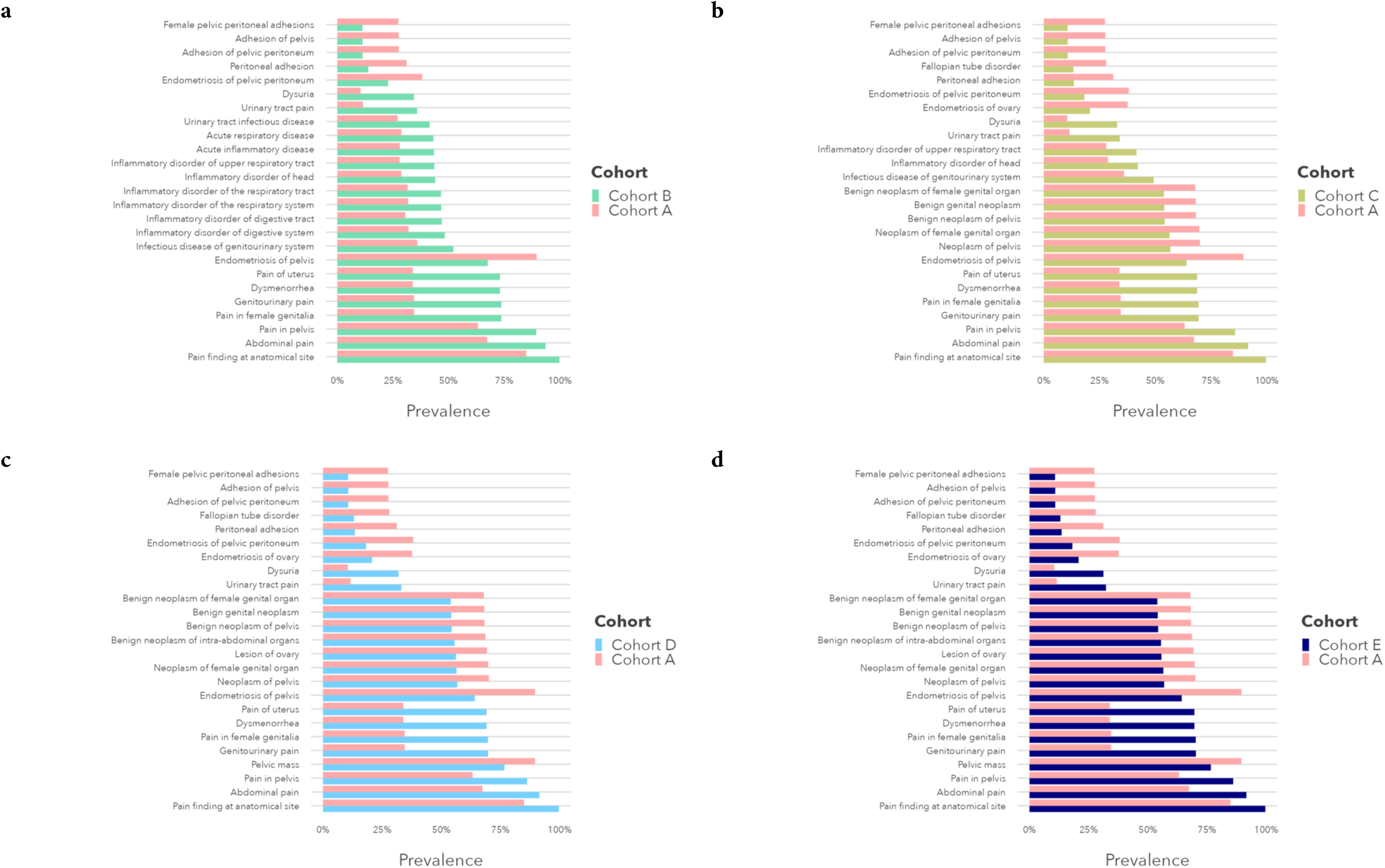
Condition Concepts in the CUIMC EHR Dataset with the Largest Differences in Prevalence Between Endometriosis Cohorts Based in Surgical Diagnosis (Cohort A) and Symptoms-based Diagnosis (Cohorts B-E). Cohort A: Surgical confirmation phenotype; Cohort B: Imaging and guideline-recognized symptoms phenotype; Cohort C: Guideline-recognized symptoms only phenotype; Cohort D: Guideline-recognized symptoms and/or pelvic pain phenotype; Cohort E: Guideline-recognized symptoms, pelvic pain, and/or abdominal pain phenotype. *Abbreviations:* CUIMC, Columbia University Irving Medical Center. EHR, electronic health record.

**Figure S15a-b.**
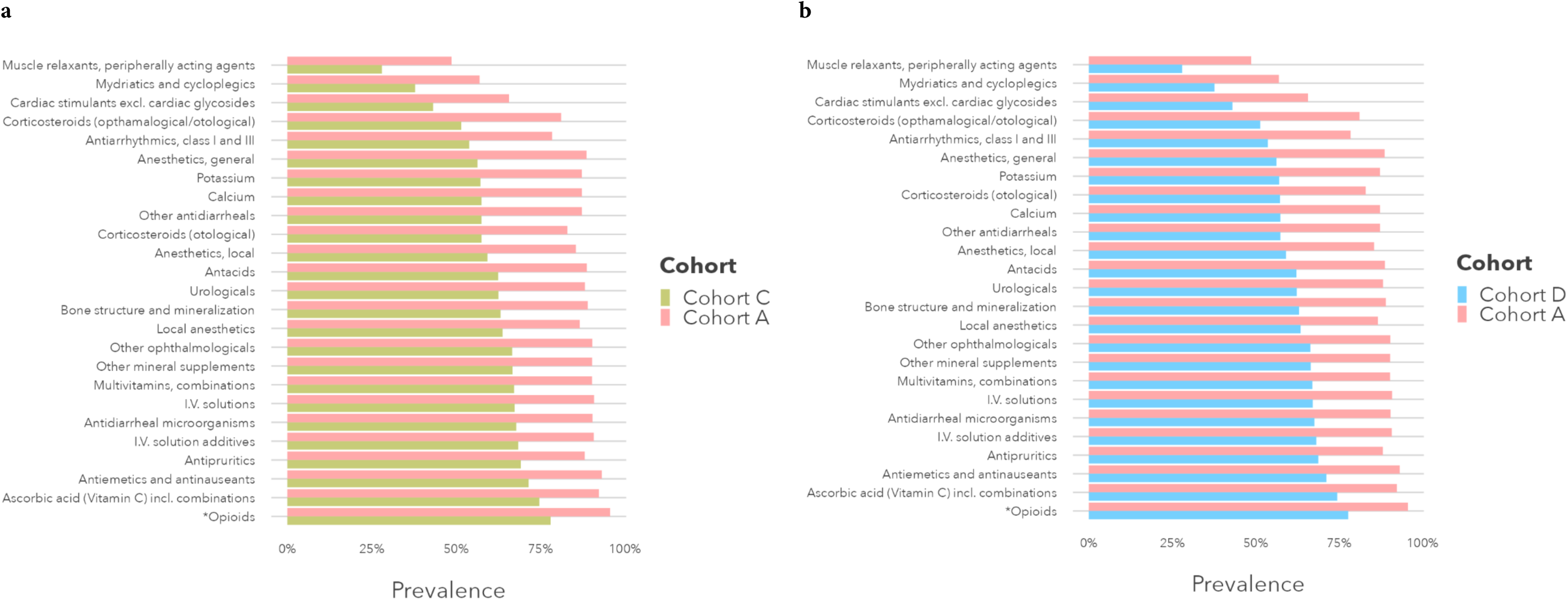
Medication Concepts in the Optum EHR Dataset with the Largest Differences in Prevalence between Endometriosis Cohorts Based on Surgical Diagnosis (Cohort A) and Imaging and/or Symptoms-based Diagnosis (Cohorts C and D). Nonsteroidal anti-inflammatory and antirheumatics, other analgesics, corticosteroids, and drugs for gastrointestinal disorders were more common in Cohort A. Hormones, antidepressants, anxiolytics, and antibacterials were more common in Cohorts C and D. *Denotes medications included among ESHRE treatment endometriosis guidelines. Cohort A: Surgical confirmation phenotype; Cohort C: Guideline-recognized symptoms only phenotype; Cohort D: Guideline-recognized symptoms and/or pelvic pain phenotype. *Abbreviations:* EHR, electronic health record.

**Figure S16a-d.**
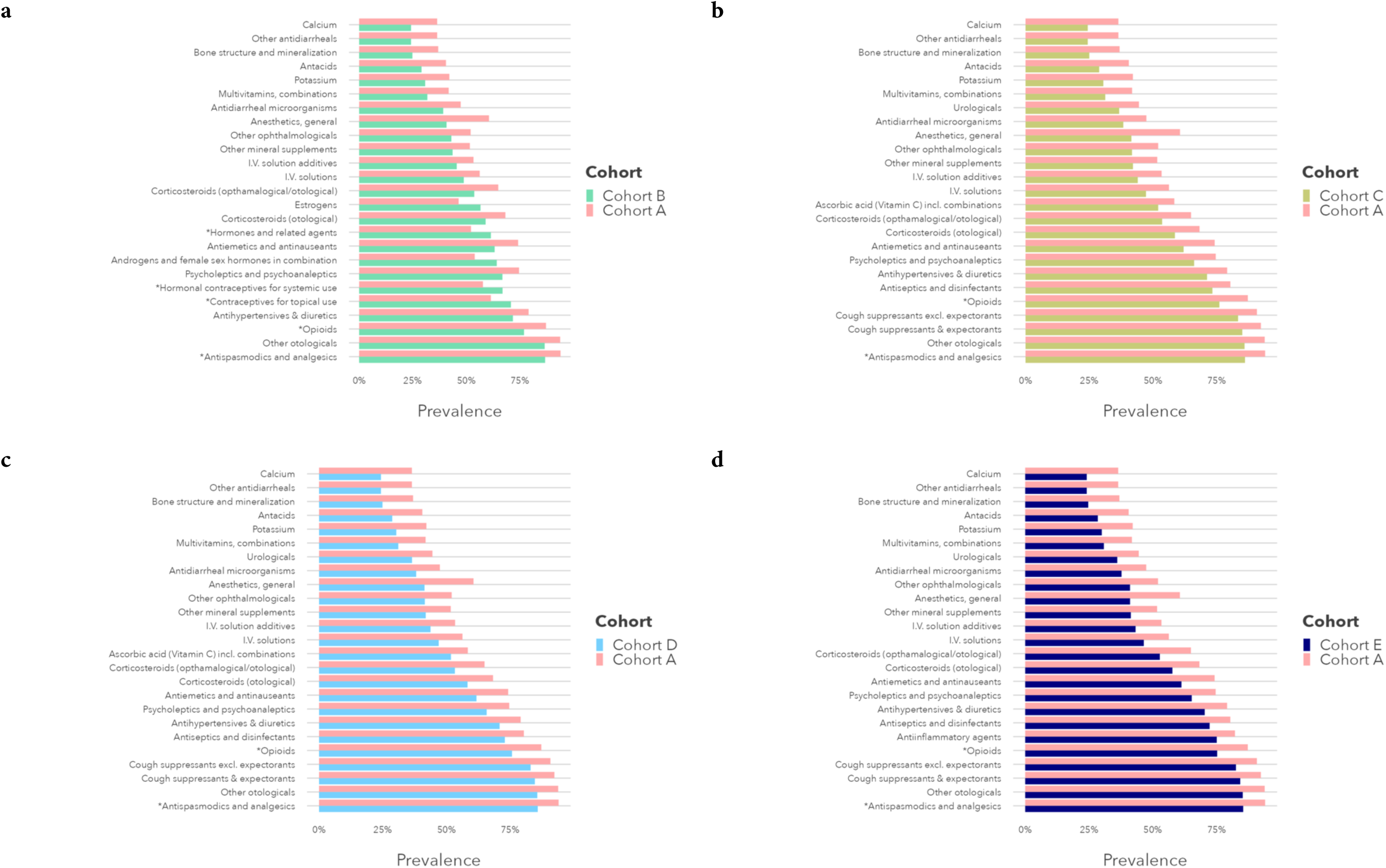
Medication Concepts in the CCAE Dataset with the Largest Differences in Prevalence Between Endometriosis Cohorts Based in Surgical Diagnosis (Cohort A) and Symptoms-based Diagnosis (Cohorts B-E). *Denotes medications included among ESHRE treatment endometriosis guidelines. Cohort A: Surgical confirmation phenotype; Cohort B: Imaging and guideline-recognized symptoms phenotype; Cohort C: Guideline-recognized symptoms only phenotype; Cohort D: Guideline-recognized symptoms and/or pelvic pain phenotype; Cohort E: Guideline-recognized symptoms, pelvic pain, and/or abdominal pain phenotype. *Abbreviations:* CCAE, Commercial Claims and Encounters.

**Figure S17a-d.**
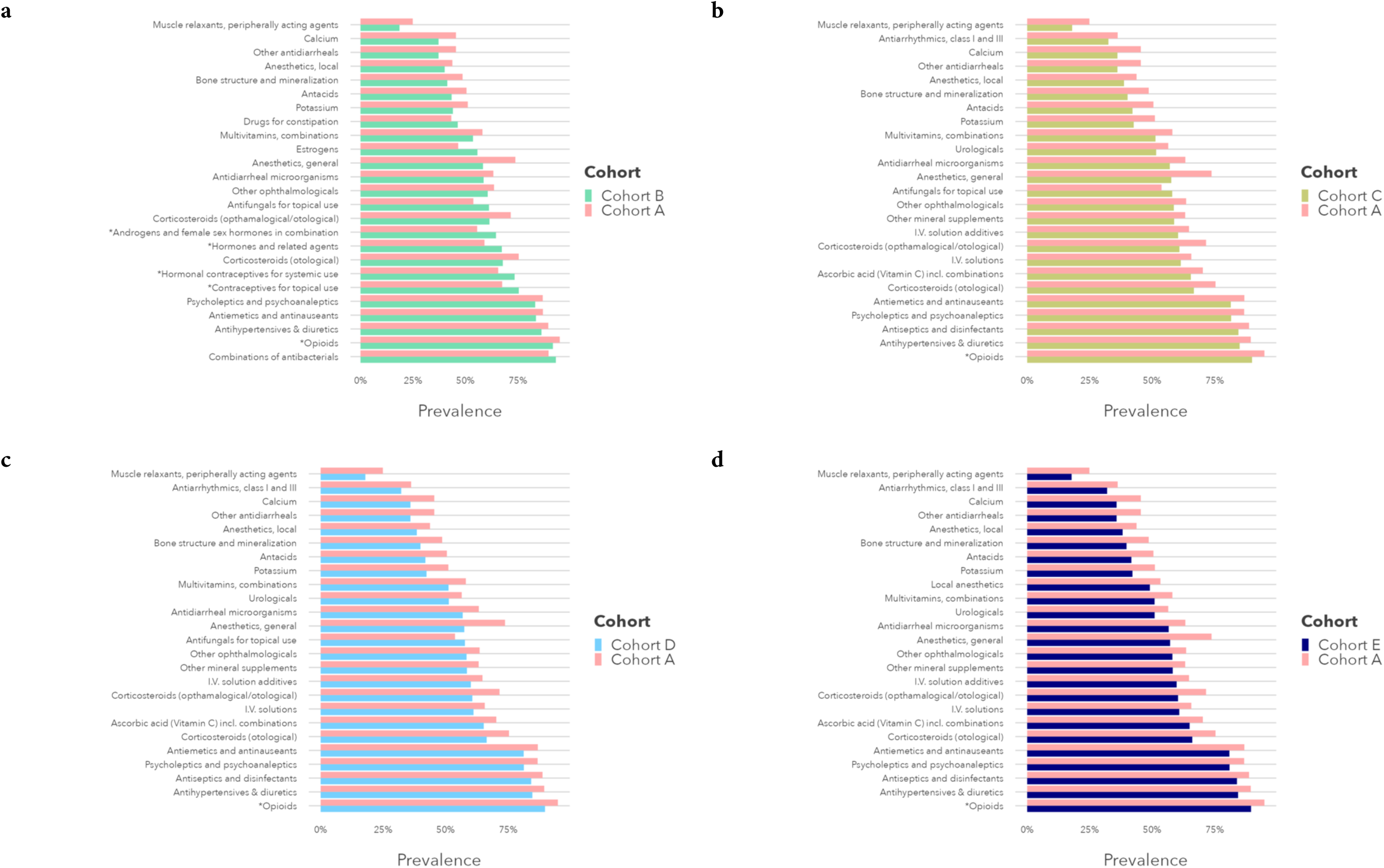
Medication Concepts in the MDCD Dataset with the Largest Differences in Prevalence Between Endometriosis Cohorts Based in Surgical Diagnosis (Cohort A) and Symptoms-based Diagnosis (Cohorts B-E). *Denotes medications included among ESHRE treatment endometriosis guidelines. Cohort A: Surgical confirmation phenotype; Cohort B: Imaging and guideline-recognized symptoms phenotype; Cohort C: Guideline-recognized symptoms only phenotype; Cohort D: Guideline-recognized symptoms and/or pelvic pain phenotype; Cohort E: Guideline-recognized symptoms, pelvic pain, and/or abdominal pain phenotype. *Abbreviations:* MDCD, Medicaid.

**Figure S18a-d.**
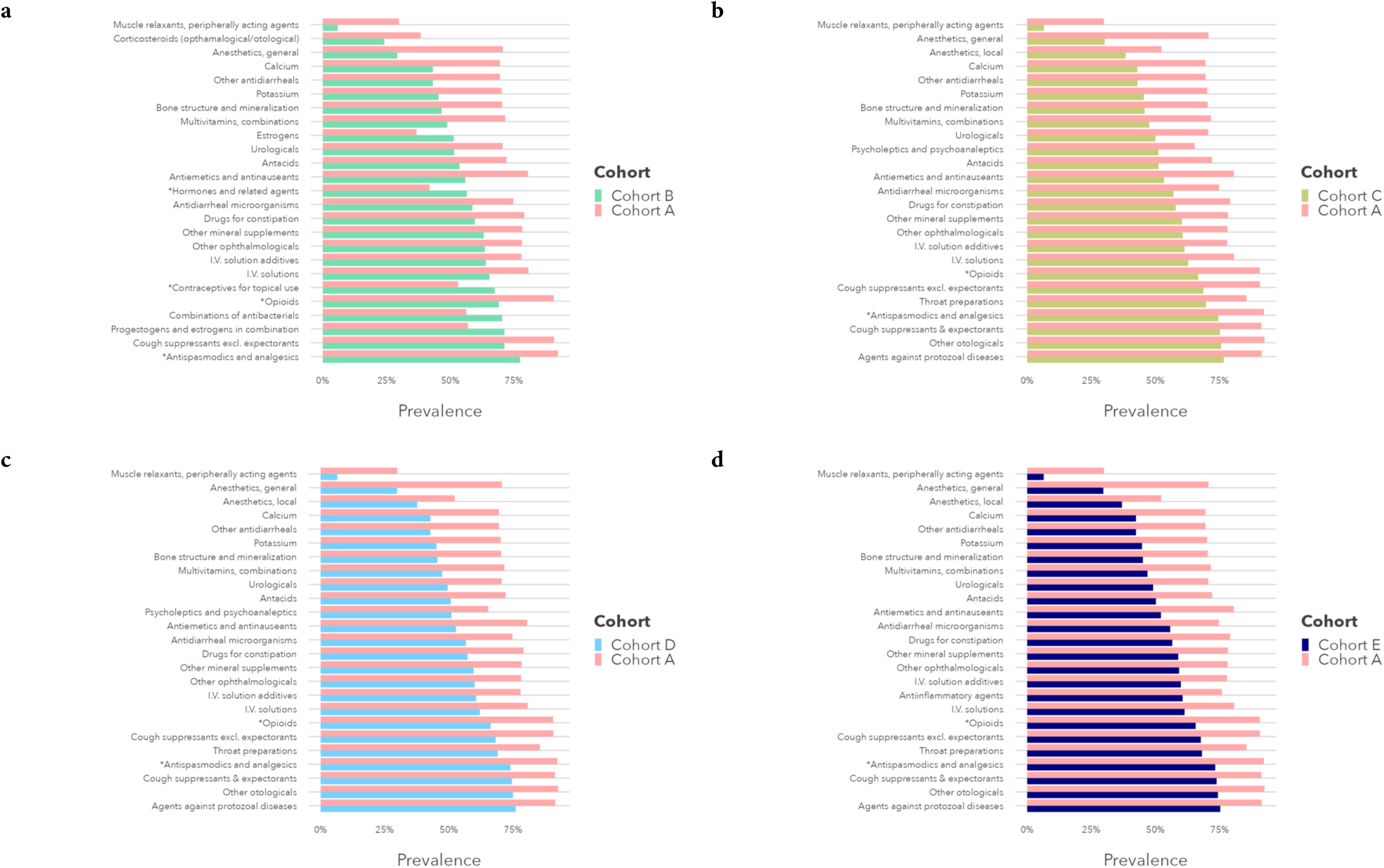
Medication Concepts in the CUIMC EHR Dataset with the Largest Differences in Prevalence Between Endometriosis Cohorts Based in Surgical Diagnosis (Cohort A) and Symptoms-based Diagnosis (Cohorts B-E). *Denotes medications included among ESHRE treatment endometriosis guidelines. Cohort A: Surgical confirmation phenotype; Cohort B: Imaging and guideline-recognized symptoms phenotype; Cohort C: Guideline-recognized symptoms only phenotype; Cohort D: Guideline-recognized symptoms and/or pelvic pain phenotype; Cohort E: Guideline-recognized symptoms, pelvic pain, and/or abdominal pain phenotype. *Abbreviations:* CUIMC, Columbia University Irving Medical Center. EHR, electronic health record.

**Figure S19a-b.**
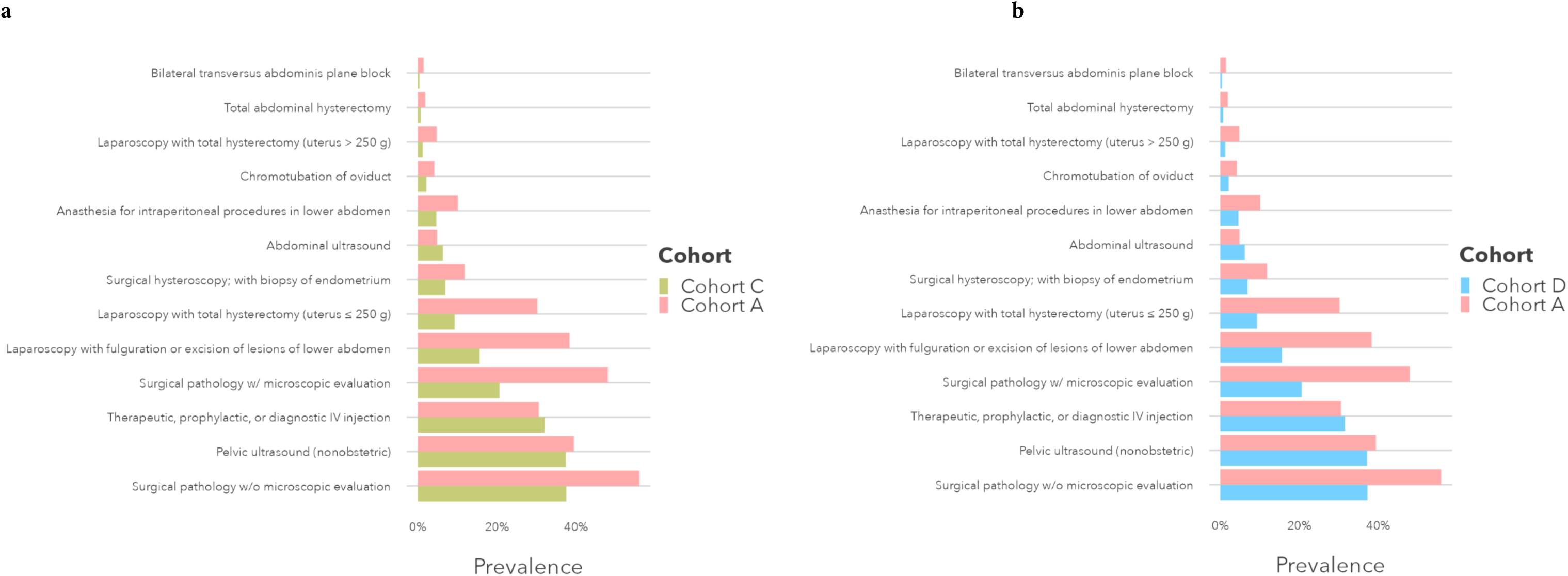
Procedure Concepts in the Optum EHR Dataset with the Largest Differences in Prevalence between Endometriosis Cohorts Based on Surgical Diagnosis (Cohort A) and Imaging and/or Symptoms-based Diagnosis (Cohorts C and D). Cohort A: Surgical confirmation phenotype; Cohort C: Guideline-recognized symptoms only phenotype; Cohort D: Guideline-recognized symptoms and/or pelvic pain phenotype. *Abbreviations:* EHR, electronic health record.

**Figure S20a-d.**
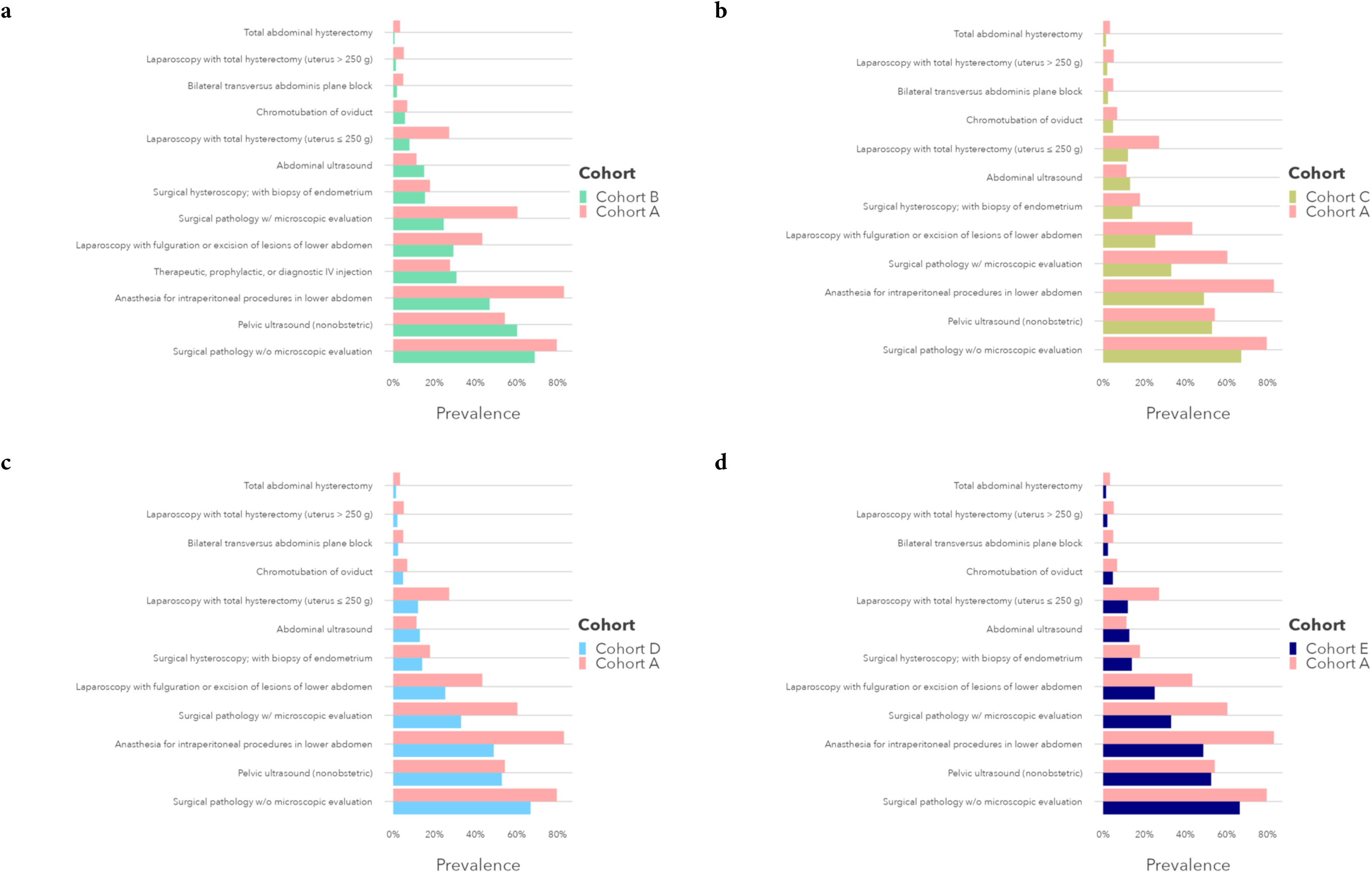
Procedure Concepts in the CCAE Dataset with the Largest Differences in Prevalence Between Endometriosis Cohorts Based in Surgical Diagnosis (Cohort A) and Symptoms-based Diagnosis (Cohorts B-E). Cohort A: Surgical confirmation phenotype; Cohort B: Imaging and guideline-recognized symptoms phenotype; Cohort C: Guideline-recognized symptoms only phenotype; Cohort D: Guideline-recognized symptoms and/or pelvic pain phenotype; Cohort E: Guideline-recognized symptoms, pelvic pain, and/or abdominal pain phenotype. *Abbreviations:* CCAE, Commercial Claims and Encounters.

**Figure S21a-d.**
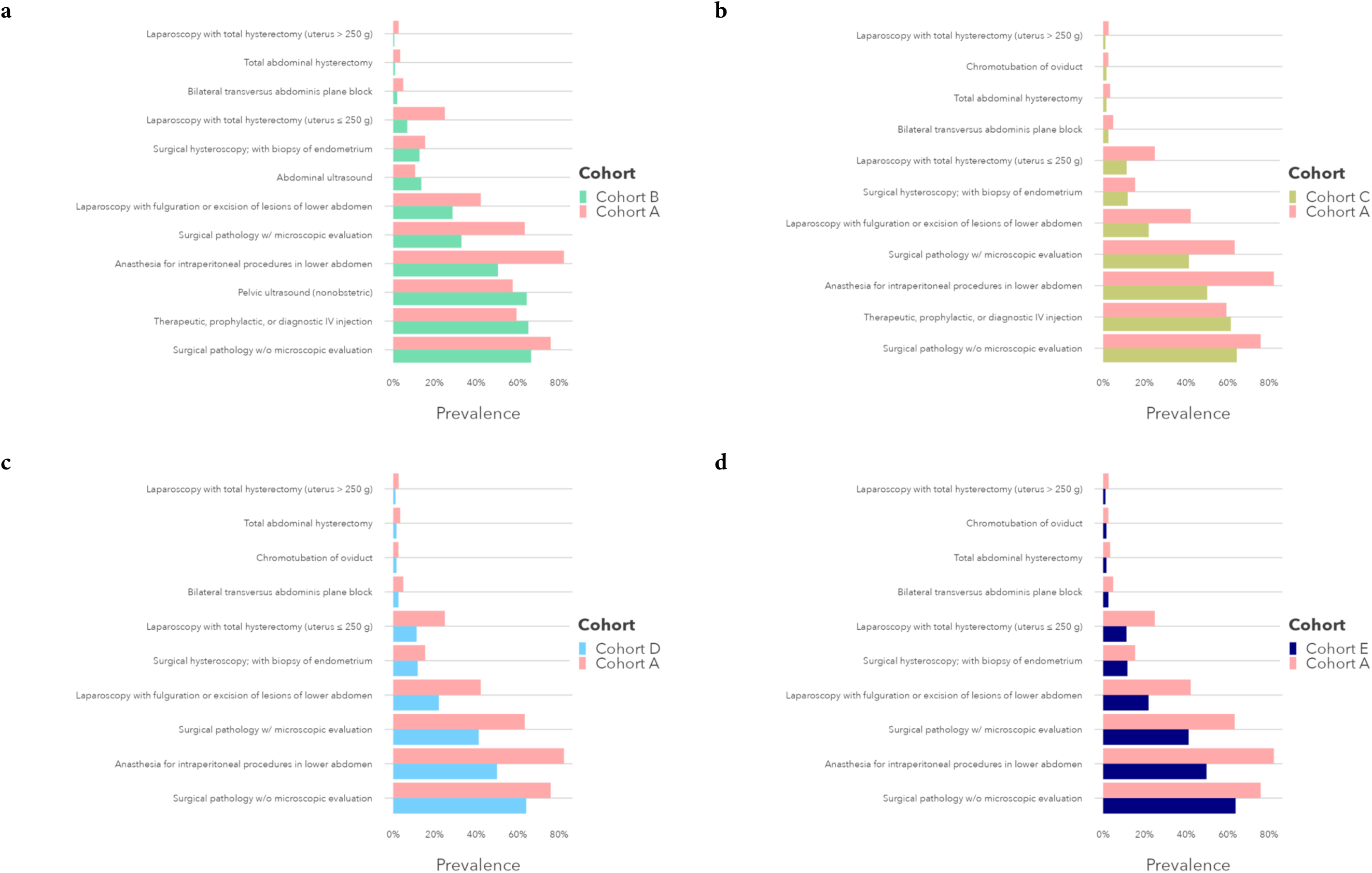
Procedure Concepts in the MDCD Dataset with the Largest Differences in Prevalence Between Endometriosis Cohorts Based in Surgical Diagnosis (Cohort A) and Symptoms-based Diagnosis (Cohorts B-E). Cohort A: Surgical confirmation phenotype; Cohort B: Imaging and guideline-recognized symptoms phenotype; Cohort C: Guideline-recognized symptoms only phenotype; Cohort D: Guideline-recognized symptoms and/or pelvic pain phenotype; Cohort E: Guideline-recognized symptoms, pelvic pain, and/or abdominal pain phenotype. *Abbreviations:* MDCD, Medicaid.

**Figure S22a-d.**
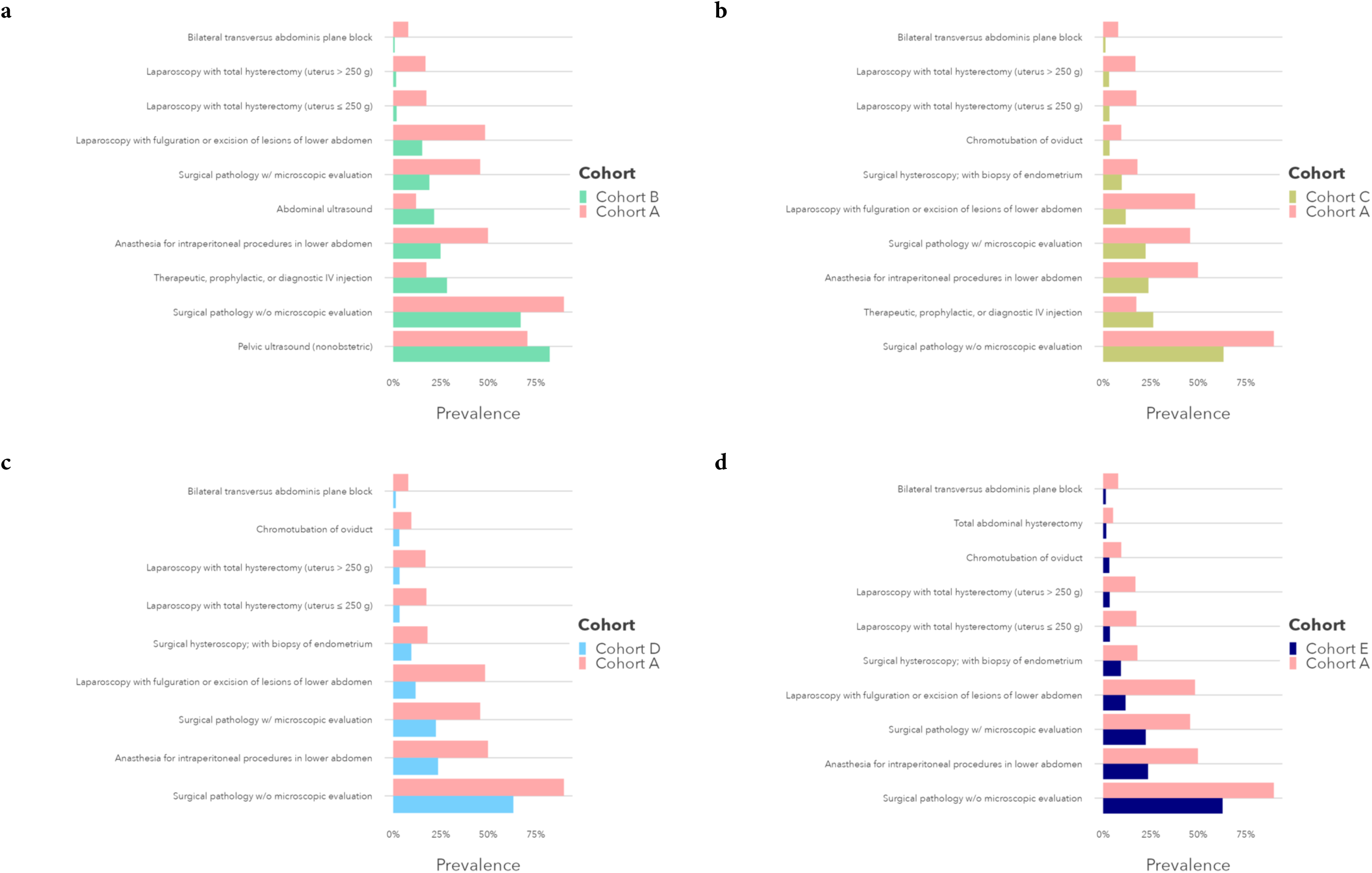
Procedure Concepts in the CUIMC EHR Dataset with the Largest Differences in Prevalence Between Endometriosis Cohorts Based in Surgical Diagnosis (Cohort A) and Symptoms-based Diagnosis (Cohorts B-E). Cohort A: Surgical confirmation phenotype; Cohort B: Imaging and guideline-recognized symptoms phenotype; Cohort C: Guideline-recognized symptoms only phenotype; Cohort D: Guideline-recognized symptoms and/or pelvic pain phenotype; Cohort E: Guideline-recognized symptoms, pelvic pain, and/or abdominal pain phenotype. *Abbreviations:* CUIMC, Columbia University Irving Medical Center. EHR, electronic health record.

